# Transferability of genetic loci and polygenic scores for cardiometabolic traits in British Pakistanis and Bangladeshis

**DOI:** 10.1101/2021.06.22.21259323

**Authors:** Qin Qin Huang, Neneh Sallah, Diana Dunca, Bhavi Trivedi, Karen A. Hunt, Sam Hodgson, Samuel A. Lambert, Elena Arciero, Genes & Health Research team, John Wright, Chris Griffiths, Richard C. Trembath, Harry Hemingway, Michael Inouye, Sarah Finer, David A. van Heel, Thomas Lumbers, Hilary C. Martin, Karoline Kuchenbaecker

## Abstract

**Background:** Individuals with South Asian ancestry have higher risk of heart disease than other groups in Western countries; however, most genetic research has focused on European-ancestry (EUR) individuals. It is unknown whether reported genetic loci and polygenic scores (PGSs) for cardiometabolic traits are transferable to South Asians, and whether PGSs have utility in clinical settings.

**Methods:** Using data from 22,000 British Pakistani and Bangladeshi individuals with linked electronic health records from the Genes & Health cohort (G&H), we conducted genome-wide association studies (GWAS) and characterised the genetic architecture of coronary artery disease (CAD), body mass index (BMI), lipid biomarkers and blood pressure. We applied a new technique to assess the extent to which loci from GWAS in EUR samples were transferable. We tested how well existing findings from EUR studies performed in genetic risk prediction and Mendelian randomisation in G&H.

**Results:** Trans-ancestry genetic correlations between G&H and EUR samples for the tested traits were not significantly lower than 1, except for BMI (r_g_=0.85, p=0.02). We found evidence for transferability for the vast majority of loci from EUR discovery studies that were sufficiently powered to replicate in G&H. PGSs showed variable transferability in G&H, with the relative accuracy compared to EUR (ratio of incremental r^2^/AUC) ≥0.95 for HDL-C, triglycerides, and blood pressure, but lower for BMI (0.78) and CAD (0.42). We observed significant improvement in categorical net reclassification in G&H (NRI=3.9%; 95% CI 0.9–7.0) when adding a previously developed CAD PGS to clinical risk factors (QRISK3). We used transferable loci as genetic instruments in trans-ancestry Mendelian randomisation and found evidence of an increased CAD risk for higher LDL-C and BMI, and for lower HDL-C in G&H, consistent with our findings for EUR samples.

**Conclusions:** The genetic loci for CAD and its risk factors are largely transferable from EUR studies to British Pakistanis and Bangladeshis, whereas the transferability of PGSs varies greatly between traits. Our analyses suggest clinical utility for addition of PGS to existing clinical risk prediction tools for this population.

**Clinical Perspective:** *What is new?:*

- This is the first study to explore the transferability of GWAS findings and PGSs for CAD and related cardiometabolic traits in British Pakistani and Bangladeshi individuals from a cohort with real-world electronic clinical data.
- We propose a new approach to assessing transferability of GWAS loci between populations, which can serve as a new methodological standard in this developing field.
- We find evidence of overall high transferability of GWAS loci in British Pakistanis and Bangladeshis. BMI, lipids and blood pressure show the highest transferability of loci, and CAD the lowest.
- The transferability of PGSs varied between traits, being high for HDL-C, triglycerides and blood pressure but more modest for CAD, BMI and LDL-C.
- Our results suggest that, for some traits, the use of transferable GWAS loci improves the robustness of Mendelian randomisation estimates in non-Europeans.

*What are the clinical implications?:*

- The polygenic score for CAD derived from genetic studies of European individuals improves reclassification on top of clinical risk factors in British Pakistanis and Bangladeshis. The improvement was driven by identification of more cases in younger individuals (25–54 years old), and of controls in older individuals (55–84 years old).
- Incorporation of the polygenic score for CAD into risk prediction models is likely to prevent cardiovascular events and deaths in this population.

## Introduction

Individuals with South Asian ancestry (SAS) account for more than a fifth of the global population and experience a higher risk of coronary artery disease (CAD) than other ancestries. For example, British South Asians have three-to four-fold higher CAD risk than White British people ^1^. Understanding the determinants of excess CAD burden in SAS populations and improving prediction to enable preventive interventions represent important public health priorities.

Common genetic variation is an important determinant of CAD and of upstream risk factors such as blood pressure, lipids, and body mass index (BMI). The genetic component of disease risk can be harnessed to identify underlying disease genes and pathways, to estimate the unconfounded effects of risk factors by Mendelian randomisation (MR), and to improve risk prediction through the application of polygenic scores (PGS). However, the genetic basis of CAD risk is not well characterised in SAS populations because genome-wide association studies (GWAS) have been mostly limited to European-ancestry (EUR) populations ^2^.

Fundamental questions remain about the extent to which the genetic determinants of cardiometabolic traits are shared by EUR and SAS populations. These have important implications to translational applications of genetic data such as causal inference with MR which could prioritise different prevention strategies or drug targets between ancestries, and clinical risk prediction. Whilst the predictive performance of PGSs derived from EUR populations in non-EUR individuals decreases with genetic distance ^3–6^, the extent to which this attenuation is due to genetic drift (differences in linkage disequilibrium and allele frequency ^7^) *versus* heterogeneity of causal genetic effects remains unclear. Furthermore, the potential clinical utility of a CAD PGS in a real-world healthcare system is largely unknown, since previous studies have mostly examined research cohorts composed of volunteers who are healthier and wealthier than average (e.g. UK Biobank ^8–12^).

Here, we perform a comparative analysis of the genetics of CAD and upstream cardiometabolic traits in EUR and SAS populations, using data from the Genes & Health (G&H) cohort ^13^. G&H is a community-based cohort of British Pakistani and Bangladeshi (BPB) individuals with linked electronic health record (EHR) data (N=22,490 individuals). This unique cohort represents an understudied and clinically vulnerable population with high levels of socioeconomic deprivation, and this is the first major genetic study focused on it. We apply new approaches to the replication of genomic risk loci across populations, perform ancestry-specific and trans-ancestry MR analysis, investigate the transportability of PGSs for CAD and its risk factors, and estimate the incremental improvement in CAD prediction when incorporating the CAD PGS into clinical risk tools.

## Methods

### Genes & Health cohort

Genes & Health (G&H) is a community-based cohort of BPB individuals recruited primarily in East London ^13^. All participants have consented for lifelong EHR access and genetic analysis. The study was approved by the London South East NRES Committee of the Health Research Authority (14/LO/1240). 97.4% of participants in G&H are in the lowest two quintiles of the Index of Multiple Deprivation in the UK. About two thirds are British Bangladeshi and the remainder British Pakistani. The median age at recruitment was 37 and 43 years for female and male participants, respectively (**Figure S1**). The cohort is broadly representative of the background population with regard to age, but slightly over-sampled females and those with medical problems since two-thirds of people were recruited in healthcare settings such as GP surgeries ^13^. We used the 2020 February data release which contained 28,022 individuals genotyped on the Illumina Infinium Global Screening Array v3 chip (with the additional multi-disease variants). Of these, 22,490 (80%) individuals had linkage to primary or secondary care data, of which 56.5% were female. Having identified related individuals (second degree or closer; kinship coefficient >0.0884) using KING v2.2.4^14^, we performed principal component analysis (PCA) in unrelated samples, and projected the remainder onto the same PC space using smartpca from EIGENSOFT v7.2.1^15^.

### Quality control and imputation of genotype data from Genes & Health

Quality control of genotype data was performed using Illumina’s GenomeStudio and plink v1.9. We first removed variants with cluster separation scores <0.57, Gentrain score <0.7, excess of heterozygotes >0.03, or ChiTest 100 (Hardy-Weinberg test) <0.6 in GenomeStudio, as well as variants that were included on the array in order to tag specific structural variants. We removed samples with low call rate (<0.995 for male samples and <0.992 for female samples across all 637,829 variants including those on Y chromosome for males) and those that failed gender checks. When there were duplicate samples, we retained the sample with the highest call rate. Using plink, we further removed variants with low call rate (<0.99), and the variant with the lowest call rate amongst duplicate variant pairs. We excluded rare variants with minor allele frequency (MAF) <1%. The high levels of autozygosity in this cohort can cause variants to fail Hardy-Weinberg equilibrium test. We thus removed variants that failed the Hardy-Weinberg test (p<1×10^−6^) in a subset of samples with low level of autozygosity. To define these ‘low-autozygosity’ individuals, we pruned SNPs (LD r^2^ >0.8) and called runs of homozygosity (RoHs) using plink1.9 with default parameters, then took the 64% of the individuals who had a fraction of the genome in RoHs <0.5%. We excluded individuals who did not have Bangladeshi or Pakistani ancestry (further than +/- 3 standard deviations [SD] from the mean of PC1 for the individuals who self-reported as coming from that group), and those who self-reported as coming from other ethnic groups or who did not report this information (**Figure S2**).

We used the Michigan Imputation Server^16^ to perform imputation with the GenomeAsia pilot reference panel^17^, imputing from 336,133 autosomal, biallelic SNPs with matched alleles. Eagle v2.4 and Minimac v4 were used for phasing and imputation, respectively. We excluded SNPs with imputation INFO score <0.3 or MAF <0.1%, which left 9,527,863 autosomal SNPs.

### Quality control and imputation of genotype data from eMERGE

We used EUR samples from the eMERGE cohort (henceforth eMERGE), a consortium of US medical research institutions, to compare with BPB individuals from G&H. Network Phase III data (N=61,377) were downloaded from dbGaP (Accession number: phs001584.v1.p1). Quality control of genotype data and imputation to the Human Reference Consortium (HRC) reference panel have been described previously ^18^. To identify EUR samples, we performed PCA in samples from the 1000 Genomes project phase 3 dataset, and projected eMERGE participants onto the same PC space using smartpca from EIGENSOFT v7.2.1^15^. For PCA, we restricted to LD-pruned common SNPs (MAF ≥1%) with imputation INFO score ≥0.98 in eMERGE. We identified samples that were clustered together with the EUR samples from the 1000 Genomes project using a dimension reduction method, Uniform Manifold Approximation and Projection (UMAP), applied to the first 20 PCs, performed using the R package “umap” v0.2.6.0^19^. Self-reported Hispanic or Latino, African, Asian, American Indian or Alaska Native individuals were excluded. This resulted in 43,877 EUR individuals available for the comparison with G&H. Well-imputed (INFO ≥0.3) bi-allelic SNPs with MAF ≥0.1% (N=11,625,805) were retained for downstream analysis.

### Phenotype and covariate definitions from electronic health-record data in Genes & Health

Of the 22,490 genotyped G&H individuals with EHR data, 20,830 had primary care data available through the Discovery Data Service^20^ which includes clinical observations as well as current and historic diagnoses (coded using READ version 2 codes, and recently converted to SNOMED CT codes using standard mapping protocols^21^). 17,226 had diagnosis and procedure codes (ICD10 and OPCS4 codes, respectively) extracted from the UK’s largest secondary care health provider, Barts Health NHS Trust.

Coronary artery disease (CAD) cases and controls were defined using the same ICD10 and OPCS4 codes as Khera *et al*. ^22^ (**Table S1**). We defined CAD cases as those with myocardial infarction or coronary revascularization in either primary and secondary care data. We excluded individuals with angina, chronic ischemic heart disease, aneurysm or atherosclerotic cardiovascular disease from the control sample ^23^. Since procedure codes were not available in eMERGE, we performed a sensitivity analysis in G&H to investigate the effects of excluding OPCS4 codes in CAD ascertainment. For this, we defined CAD solely using ICD10 codes in individuals with secondary care data, ignoring OPCS codes and primary care data; we excluded individuals without secondary care data for this analysis.

We used median adult height and weight measurements within the past 5 years to calculate BMI. For lipids, we took the latest adult measurements and corrected for statin usage if lipid levels were measured between the start and end date of any statin prescriptions. No adjustment was made on HDL cholesterol (HDL-C) or triglycerides. Adjustment of lipids followed the procedure in Liu *et al*. ^24^, as follows. To correct for statin usage, total cholesterol (TC) was replaced by TC/0.8. LDL cholesterol (LDL-C) levels were calculated using the Friedewald equation, and statin-adjusted LDL-C was recalculated using adjusted TC levels as follows: corrected LDL-C = uncorrected LDL-C + 0.2*adjusted TC. LDL-C/0.7 was used for 32 individuals for whom we couldn’t find a TC measurement on the same date. Rank-based inverse normal transformation was applied to the lipid levels.

We extracted the latest systolic blood pressure (SBP) and diastolic blood pressure (DBP) measurements and adjusted for blood pressure medication use by adding 15 and 10 mmHg to SBP and DBP, respectively, if the measurement coincided with any prescription date ^25^. Sample sizes are shown in **Table 1** (all individuals) and **Table S2** (unrelated).

**Table 1.** Reproducibility of loci for cardiometabolic phenotypes in British Pakistani and Bangladeshis. Note that when assessing sharing of causal variants, we excluded loci where the overlap between UKBB and G&H was <10 SNPs and SNP coverage of the region was low (<10%). *For SBP and DBP, power was calculated with observed effect size in normalised BP values.

| Trait | N. samples<br>(cases:controls) | N.<br>established loci | N. observed<br>transferable<br>loci (%) | Percentage<br>of expected<br>transferable<br>loci | Observed/<br>Expected | N. transferable<br>loci with shared<br>causal variant/N.<br>transferable loci<br>assessed (%) |
| --- | --- | --- | --- | --- | --- | --- |
| **BMI** | 16890 | 662 | 140 (21%) | 20% | 1.05 | 15/58 (26%) |
| **LDL-C** | 12746 | 82 | 51 (62%) | 50% | 1.24 | 15/32 (47%) |
| **HDL-C** | 14944 | 103 | 66 (64%) | 56% | 1.14 | 14/29 (48%) |
| **TC** | 15641 | 107 | 61 (57%) | 49% | 1.16 | 23/38 (61%) |
| **TG** | 13037 | 95 | 47 (49%) | 47% | 1.04 | 14/25 (56%) |
| **DBP** | 18536 | 175 | 36 (21%) | 21%* | 1.00 | NaN |
| **SBP** | 18536 | 171 | 30 (18%) | 18%* | 1.00 | NaN |
| **CAD** | 22008<br>(1110:20898) | 71 | 9 (13%) | 21% | 0.62 | NaN |

To calculate a standard clinical risk score to compare with the PGS, we calculated the QRISK3 10-year predicted risk for CAD ^26^ in G&H using the R package “QRISK3” v0.3.0 ^27^. QRISK3 was calculated based on the data available up until 1 January, 2010, which is about 10 years prior to the latest data extraction. We excluded about one third of CAD cases whose diagnosis was made earlier than this assessment date (prevalent cases) and used incident cases who developed CAD later. Follow-up varied for cases and was fixed at 10 years for controls. We used clinical data that were extracted earlier than the assessment date (1 January 2010) to calculate QRISK3. The QRISK3 algorithm has variables that indicate whether a patient has a variety of other diseases, and these were defined using the codes shown in **Table S3**, following ^10^. Medication use (hypertension treatment, corticosteroid, and atypical antipsychotic medication) was defined as two or more prescriptions, with the most recent one having been issued within 28 days prior to the assessment. We used the most recent measurements taken prior to the assessment date, and kept individuals with at least three non-missing measurements out of four (height, weight, SBP, and TC). Pattern of missingness is shown in **Figure S3**. Townsend index was not available in G&H, so we used the mean value (3.307) of the lowest two quintiles from the 2011 census data in the UK ^28^. HDL-C levels were all measured later than 2010 in G&H, so for TC/HDL-C ratio, we used 3.905 and 4.882 (averages calculated using later data) for females and males, respectively. To deal with missing data, we applied multiple imputation which accounts for sex, age, and genetically-defined ancestry (Bangladeshi *versus* Pakistani; identified using PCA-UMAP), using the R package “mice” v3.13.0 to impute height, weight, SBP, SD of SBP measurements within 2 years, and smoking status.

### Phenotype definitions from electronic health-record data in eMERGE

Phenotype data in eMERGE were downloaded from dbGaP (phs001584.v1.p1, phs000888.v1.p1, phs001584.v2.p2). Individuals younger than 16 years old were excluded. BMI was provided and we took the median value from adult measurements. Lipid and blood pressure measurements were taken from dataset phs000888.v1.p1. Data on medications affecting lipid and BP measurements were not available, so the highest measurements for LDL, TC, SBP, and DBP were used when comparing PGSs with G&H in order to minimise the effects of medications. CAD was ascertained using ICD9/10 codes which were available in the updated eMERGE Phase III dataset (phs001584.v2.p2). Coronary artery disease (CAD) cases and controls were defined based on secondary care ICD10 codes as described above for G&H (**Table S1**).

### Genome-wide association analyses in Genes & Health

GWAS was performed with SAIGE ^29^ and adjusted for age, age^2^, sex and the first twenty principal components. For total cholesterol and LDL-C, adjustments were made for use of statins as described above. We followed the QC procedure in ^30^ (*EasyQC* package) with the following exclusion criteria for variants: monomorphic variants, missing / invalid estimates, allele mismatch and allele frequency difference of >0.2 with reference panel, imputation INFO score <0.7 (<0.9 for downstream analysis i.e. correlation and colocalisation), MAF <0.005 (<0.01 for downstream analysis i.e. correlation and colocalisation).

### Heritability and trans-ancestry correlations

Datasets that were used in analyses are provided in **Table S4**. We used GCTA to estimate SNP heritability in G&H and eMERGE^31^. We excluded one sample in each pair of 3^rd^-degree relatives (kinship coefficient >0.0442 calculated using KING v2.2.4^14^). We used SNPs with INFO >0.9 and MAF >0.01 in each cohort separately. We also calculated SNP heritability using the intersection of these SNP sets in both cohorts. For CAD, we estimated SNP heritability on the liability scale using 6.7% as the prevalence estimate in the US ^32^, and 3.33% for the UK background population from which G&H is sampled, defined as all people from South Asian ethnicities (N=255,066 aged ≥20 years) registered with a primary health physician/GP in four east London boroughs.

For the genetic correlation analyses, we used GWAS summary statistics generated in EUR individuals from UK Biobank (UKBB), since we needed a larger sample size of ancestrally homogeneous individuals than is available through eMERGE to obtain accurate estimates. We used Popcorn (https://github.com/brielin/Popcorn) to estimate the trans-ancestry genetic correlations between G&H and UKBB EUR individuals while accounting for differences in LD structure ^33^ (i.e. the correlation of causal-variant effect sizes across the genome at SNPs common to both populations). Variant LD scores were estimated for ancestry-matched 1000 Genomes v3 data for each study combination (i.e. SAS-EUR). The estimation of LD scores failed for chromosome 6 for some groups, so we left out the major histocompatibility complex (MHC) region (positions 28,477,797 to 33,448,354) from chromosome 6 from all comparisons. Variants with INFO score <0.9 or MAF <0.01 were excluded. A p-value <0.05 indicated that the genetic correlation was significantly less than 1 i.e. r_g_ <1.0.

### Assessment of transferability of established loci

Previous studies that evaluated reproducibility of GWAS loci in SAS individuals did not formally account for differences in power or LD patterns ^34–36^. We assessed whether established trait-associated loci were reproducible in G&H by performing a lookup of loci identified in non-SAS ancestry GWAS (**Table S4**). Credible sets for established loci were generated and consisted of lead (independent) variant plus proxy SNPs (r^2^ >=0.8) within a 50kb window (based on the EUR 1000 Genomes data) of the sentinel variant and with p-value <100 × p_sentinel_. The locus was defined as being ‘transferable’ if at least one variant from the credible set was associated at p <0.05 with the relevant trait in G&H, and the direction of effect matched in both datasets. For loci harbouring multiple signals, we only kept the most strongly associated variant (i.e. smallest p-value). Expected power for replication was calculated using alpha=0.05, the effect size estimated in the EUR GWAS, and the allele frequency of the variant and sample size in G&H. The power of lead variants per locus was summed up and divided by the number of loci to give an estimate of the number of expected significant loci per trait, which was compared with the observed number of such loci; to our knowledge, this is a novel approach for assessing reproducibility of GWAS findings. Loci were only deemed to be ‘non-transferable’ if they contained at least one variant in the credible set with >80% power and yet none of the variants in the credible set had p <0.05 and no variant within 50kb of locus had p <1×10^−3^ in G&H. LocusZoom (http://locuszoom.org/) was to create regional association plots.

### Trans-ancestry colocalisation

We used the Trans-ethnic colocalisation method (TEColoc) (https://github.com/KarolineKuchenbaecker/TEColoc)^37^ which tests whether a specific locus has the same causal variant in two groups with different ancestry, and applied it to G&H and UKBB EUR individuals. This method adopts the joint likelihood mapping (JLIM) statistic developed by Chun and colleagues^38^ that estimates the posterior probabilities for colocalisation between GWAS signals and compares them to probabilities of distinct causal variants while explicitly accounting for LD structure. For this, LD scores were estimated using a subset of samples from the 1000 Genomes Project v3 that had matching ancestry to all Europeans for UK Biobank. For G&H we used raw genotype data and LD was estimated directly for these samples. JLIM assumes only one causal variant within a region in each study. We therefore used small windows of 50Kb for each known locus to minimise the risk of interference from additional association signals. Distinct causal variants were defined by separation in LD space by r^2^ ≥0.8 from each other. We excluded loci where the overlap between UKBB and G&H was <10 SNPs and the proportion of well-imputed SNPs overlapping between cohorts (SNP coverage) was <10%; this left no loci to consider for CAD, SBP and DBP. We used a significance threshold of p <0.05 to determine evidence of sharing. LocusZoom (http://locuszoom.org/) was to create regional association plots.

### Construction of polygenic risk scores

We evaluated the performance of PGSs in G&H and eMERGE. We first assessed PGSs that were previously constructed (mostly optimised in EUR samples) from the PGS Catalog ^39^. We restricted to 7,353,388 bi-allelic SNPs that had INFO ≥0.3 and MAF ≥0.1% in both eMERGE and G&H. Variant information in existing PGS was harmonised to GRCh37 using dbSNP mappings from Ensembl Variation and liftover. We calculated PGSs as weighted sums of imputed allele dosages using plink2.0 --score function. There were often multiple PGSs that were previously developed from different studies available for each trait, and below we report the one that had the highest accuracy in each cohort. The best PGS (defined as described in the next section of the Methods) for BMI was derived from GWAS conducted in primarily EUR samples and optimised in EUR individuals, and those for lipids and BP contained genome-wide significant variants identified in EUR GWASs. We selected different PGSs for CAD in eMERGE and G&H, with the former optimised in EUR individuals and the latter in SAS individuals; in both cases these were based on GWAS conducted in primarily EUR samples. The details of each PGS are in **Table S5**.

Next we calculated PGSs using the clumping and p-value thresholding method (C+T) and optimised PGSs in G&H and eMERGE separately. We used GWAS summary data from primarily EUR samples (**Table S4**). We used LD estimated using EUR samples (N=503) from the 1000 Genomes project for clumping using PRSice2 v2.2.11^40^. We calculated multiple scores using combinations of various LD r^2^ thresholds (0.1, 0.2, 0.5, 0.8) and p-value thresholds (5×10^−8^, 1×10^−7^, 5×10^−7^, 1×10^−6^, 5×10^−6^, 1×10^−5^, 5×10^−5^, 1×10^−4^, 5×10^−4^, 0.001, 0.005, 0.01, 0.02, 0.05, 0.08, 0.1, 0.2, 0.3, 0.4, 0.5, 0.8, 1) for each trait, and reported the PGS with the best predictive performance within each target cohort.

Lastly, we calculated meta-PGSs proposed by Marquez-Luna et al.^41^ that incorporate GWAS summary data from the target populations. We downloaded GWAS summary data that were generated in SAS samples of the UKBB from the Pan-UK Biobank website (https://pan.ukbb.broadinstitute.org), and constructed scores (PGS_SAS_) using the C+T method described above and using SAS samples from the 1000 Genomes project for the LD reference. We combined the scores derived from EUR GWASs (PGS_EUR_) and PGS_SAS_ in linear regression to construct meta-PGSs.

### Assessment of PGS accuracy and clinical performance

We excluded one sample in each pair of 2^nd^-degree relatives (kinship coefficient >0.0884 calculated using KING v2.2.4^14^). Individuals with the highest number of relatives (and controls, if the trait is binary) were removed first. Sample sizes for each trait are in **Table S2**. Quantitative traits were inverse normal transformed. Age at recruitment was used as a covariate for analysis of disease status, and age at measurement for analysis of quantitative traits. PGSs were standardised to a mean of 0 and SD of 1. We fitted the following two models: (1) the full model which had PGS and covariates namely sex, age, age^2^, and the first 10 genetic PCs, and (2) the reference model which accounted for the covariates only. For continuous risk factors, linear regression was fitted, and the gain in R^2^ when adding PGS as an additional predictor, or incremental R^2^, was calculated as the difference between the R^2^ of the full model and the reference model. Logistic regression was used to assess the associations between PGSs and CAD. The area under the receiver operating characteristic curve (AUC) was estimated for both models with the R package “pROC” v1.16.2 and incremental AUC was calculated similarly. We performed bootstrap resampling of individuals 1,000 times to estimate the 95% confidence intervals for incremental R^2^ and incremental AUC. The best PGS per trait was the one with the highest incremental R^2^ for continuous risk factors and the one with the highest incremental AUC for CAD. We estimated the effect size (or odds ratio for binary traits) per SD of PGS from the full model. Effect size or odds ratio for quintiles, and for top 10% versus middle 40-60% were reported as well. Relative accuracy was calculated as the ratio of incremental AUC (or incremental R^2^ for continuous traits) in G&H to that in eMERGE.

QRISK3 scores were calculated for 8,112 unrelated individuals as described above (420 CAD cases and 7,702 controls). To integrate QRISK3 scores with PGS for CAD, we followed Riveros-Mckay *et al*.^10^ and calculated an integrated score by multiplying the odds converted from the QRISK3 score with the odds ratio given an individual’s PGS, where the odds ratio per SD of PGS was estimated using a logistic regression in which QRISK3 and their interaction were accounted for. The logistic regression was performed in males and females separately. We used the most accurate PGS for CAD in SAS from the PGS Catalog, which was developed by Wang et al.^42^; this score was derived from EUR GWAS using LDpred and tuned in SAS individuals in UKBB. We regressed out 10 PCs from the PGS, and used the scaled residuals in the Cox regression analysis. Cox regression was performed using the R package “survival” v3.2-7. The concordance indices (C-indices) of the following models were compared: (1) age at assessment + gender, (2) PGS + age at assessment + gender, (3) QRISK3, and (4) the integrated score. We calculated the continuous net reclassification index (NRI) and categorical NRI (using 10% as the threshold to classify high-risk individuals) for the integrated score compared to QRISK3 alone. NRI was calculated as the sum of NRI for cases and NRI for controls (noncases):

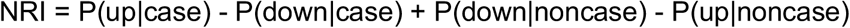

For continuous NRI, P(up|case) and P(down|case) indicate the proportions of cases that had higher or lower risk estimates using the integrated score, respectively. For categorical NRI, P(up|case) indicates the proportions of cases that were reclassified as high-risk individuals (i.e. with <10% risk by QRISK3 but >10% by the integrated scores). We calculated NRI in two age groups (25–54 versus 55–84 years old at baseline, chosen since the average age of onset in this cohort was 55.3 years old), as well as in sex-by-gender subgroups. Bootstrap resampling (1,000 times) was used to estimate confidence intervals for NRI.

### Mendelian randomisation analysis

We modelled liability to CAD as our outcome within a two-sample Mendelian randomisation^43^ (MR) framework using the risk factors (BMI, SBP, DBP, LDL-C, HDL-C, TG) as exposures. To identify genetic instruments for the exposure, we explored three alternative approaches: (a) established loci significant at p<5×10^−8^ in the original EUR GWAS; (b) transferable loci defined as described above, taking the effect size from the original EUR GWAS; and (c) loci significant at p<5×10^−8^ in the SAS ancestry group of the Pan-UKBB GWAS, LD-clumped to an r^2^<0.2 with a LD window of 50kb, based on SAS 1000 Genomes project LD reference. Where insufficient genome-wide significant instruments were identified, we used a more permissive p-value threshold of p<5×10^−5^ for instrument selection in UKBB SAS. The primary MR analysis was performed using, as outcome, summary association data from the G&H CAD GWAS performed as described above, using the inverse-variance weighted method under a random effect model, implemented with the TwoSampleMR R package^44^. For comparison, a two-sample MR approach was also performed using summary data for CAD from eMERGE and established loci significant at p<5×10^−8^ in the original EUR GWAS. We also undertook several sensitivity analyses. In brief, we evaluated the MR-Egger intercept to assess directional pleiotropy and Cochran’s Q statistic^45^ as an indicator of heterogeneity. MR analysis using weighted median^46^ and weighted methods^47^ models were additionally performed in the presence of heterogeneity.

## Results

In G&H, 4.9% (N=1,110) of the individuals had coronary artery disease (CAD), with the age of onset ranging from 17 to 97 years old (median 55). A quarter of the G&H participants were on active statin prescriptions, 23% on BP medications, 29% had high TC levels (>5 mmol/L), and 30% had high LDL-C levels (>3 mmol/L; **Table S6**) ^48^. Datasets that were used in each analysis are provided in **Table S4**.

### Shared genetic architecture of cardiometabolic traits

We compared the genetic architecture of coronary artery disease (CAD) and upstream risk factors, namely HDL-C, LDL-C, triglycerides (TG), total cholesterol (TC), systolic and diastolic blood pressure (SBP & DBP), between British Pakistanis and Bangladeshis (BPB) from G&H, and European-ancestry populations (EUR) (**Figure 1**). We used EUR individuals from the EHR-based eMERGE cohort to estimate heritability, since phenotypes had been ascertained in a similar way to G&H (i.e. EHRs). All traits were found to have significant SNP heritability (h^2^= 0.03–0.23) in G&H, with estimates similar to those in eMERGE (**Table S7, Figure 2A**), except for LDL-C, SBP and DBP which had significantly lower values in G&H than eMERGE (e.g. for LDL-C, h^2^ was 0.21 [95% CI: 0.17–0.25] in eMERGE and 0.06 [95% CI: 0.02–0.10] in G&H). Conclusions were unchanged when restricting the heritability estimates to the same set of well-imputed SNPs in both cohorts (**Table S7**). We observed high genetic correlations between G&H and EUR from UKBB for all traits, with the lowest value seen for SBP (r_g_=0.71 [95% CI: 0.36–1.06], p=0.09; **Figure 2B**). The only trait for which the genetic correlation differed significantly from one was BMI (r_g_=0.85 [95% CI: 0.71–0.99], p=0.02).

**Figure 1.**
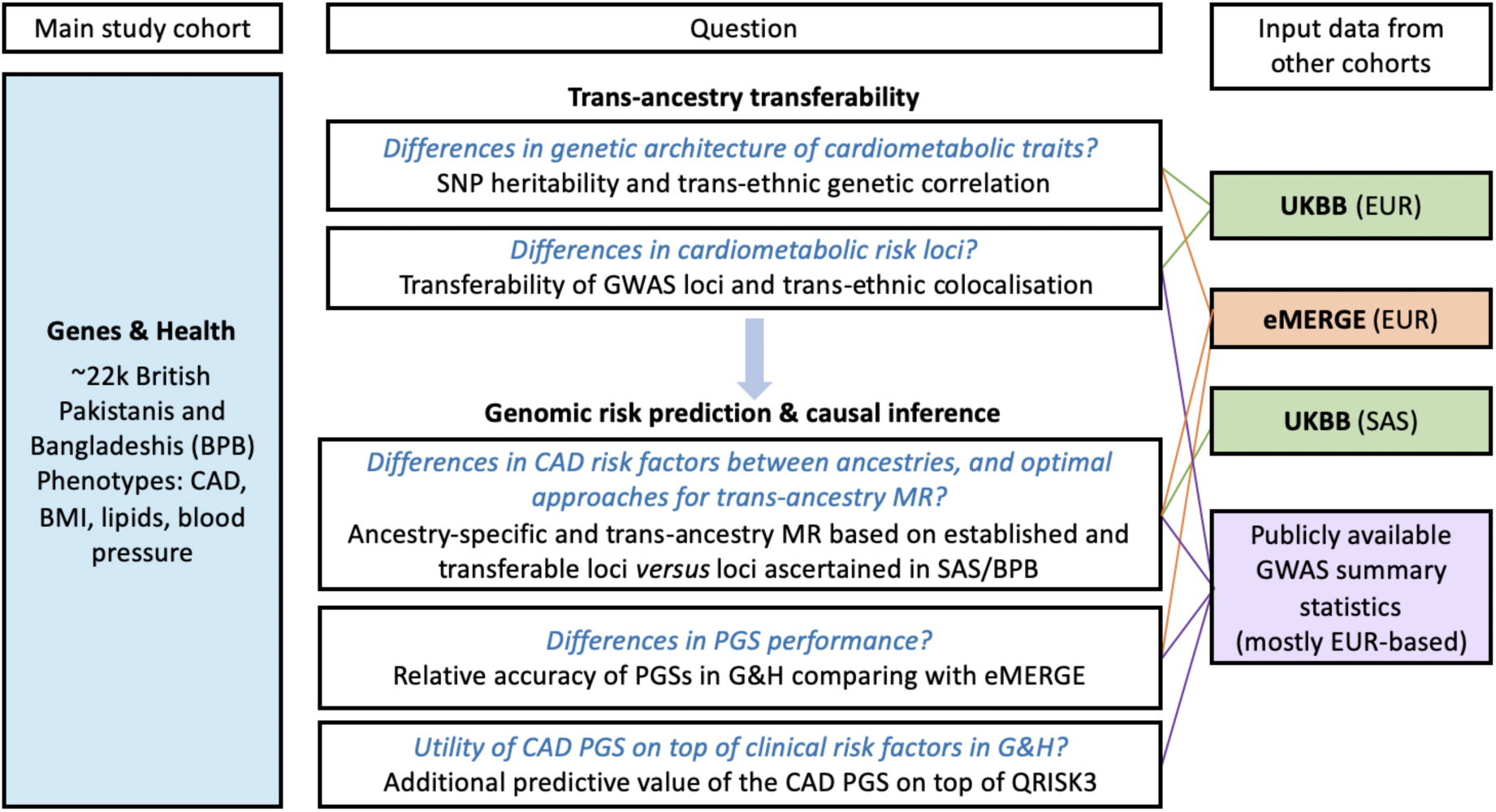
Summary of study design, research questions and analyses conducted. The coloured boxes indicate input data. Within the white boxes, black text indicates the analyses we used to address the questions in blue. BPB: British Pakistanis and Bangladeshi ancestry; EUR: European ancestry; SAS: South Asian ancestry; CAD: coronary artery disease; BMI: body mass index; SNP: single nucleotide polymorphism; GWAS: genome-wide association study; MR: Mendelian randomisation; PGS: polygenic score; UKBB: UK Biobank. Datasets and discovery GWAS that were used in each analysis are provided in **Table S4**.

**Figure 2.**
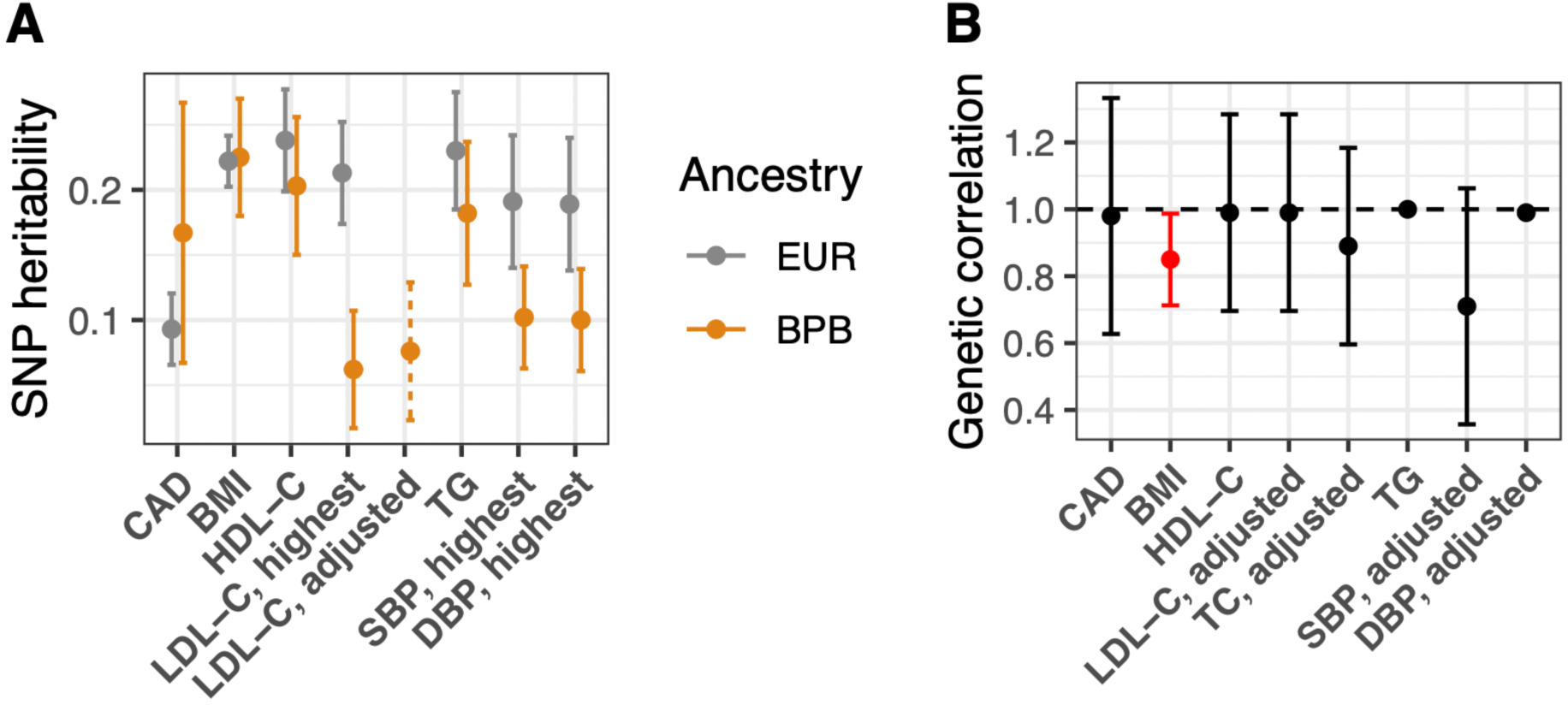
SNP heritability and trans-ancestry genetic correlations for cardiometabolic traits. **A**. SNP heritability was estimated using GCTA in G&H (orange) and eMERGE (grey) for cardiometabolic traits. Dashed line indicates statin-adjusted LDL cholesterol levels in G&H which are not available in eMERGE. Error bars represent 95% confidence intervals in both plots. **B**. Genetic correlations were estimated using GWAS summary statistics generated from G&H and European-ancestry individuals from UK Biobank. Red indicates that the genetic correlation is significantly lower than 1 (p-value = 0.02 for BMI). BMI: body-mass index; BPB: British Pakistani ancestry; CAD: coronary artery disease; EUR: European ancestry; LDL-C: low-density lipoprotein cholesterol; G&H: Genes & Health; HDL-C: high-density lipoprotein cholesterol; SBP: systolic blood pressure; DBP: diastolic blood pressure.

### High transferability of cardiometabolic loci

We assessed whether published trait-associated genomic loci identified in predominantly EUR populations were shared by the BPB population represented by G&H. To account for differences in LD patterns, our assessment of transferability was based on the credible sets of variants per locus, likely to contain the causal variant, rather than the sentinel variants alone. Low numbers of transferable loci may be due to limited statistical power rather than lack of causal variant sharing. Therefore, we compared the number of observed transferable loci with the number expected given the sample size and allele frequency in G&H if all causal variants were shared. The number of expected transferable loci varied widely between traits (e.g. we expected to be able to detect significant associations for 56% of HDL-C loci but only for 18% of SBP loci), highlighting the importance of accounting for power when assessing transferability. Across most traits examined, the observed number of transferable loci closely matched the loci we expected (**Table 1** and **Table S8**). For example, for BMI we expected to be able to find evidence for transferability for 20% of loci and we did indeed observe transferability for 21% of loci. However, the exception was CAD for which the number of observed transferable loci (13%) was below the expected number (21%), although this difference was only marginally significant (binomial p-value = 0.05).

We also assessed whether there were any specific loci that were not transferable despite being well powered to observe an association (power >80%). Out of a total of 184 well-powered loci tested across all traits, only nine were non-transferable; that is, no variant in the credible set was significant at p<0.05 and no variant within 50kb of locus was significant at p<1×10^−3^ (**Figure S4**). These nine loci were all associated with lipid traits: *EVI5, NBEAL1, GPAM, CETP, STAB1, TTC39B, SH2B3, ACP2* and *NECAP2* (**Table 2**). Of these loci, *CETP*, which was previously associated with LDL-C levels in Europeans (established variant in Europeans -rs7499892), was strongly associated with HDL-C in G&H (p=7.08×10^−56^), but not with LDL-C levels (p=0.23) (**Figure S5**) despite having >80% power for replication.

**Table 2.** Established loci from European-ancestry GWAS inferred to be non-transferable to British Pakistani and Bangladeshis. * Tag SNP: rs7499892, r^2^=1.

| Trait | SNP | Chromosome | Position | Locus | Other allele/effect allele | Effect allele frequency | p-value | Effect size |
| --- | --- | --- | --- | --- | --- | --- | --- | --- |
| LDL | rs7515577 | 1 | 93009438 | *EVI5* | C/A | 0.96 | 0.88 | 0.005 |
| LDL | rs2255141 | 10 | 113933886 | *GPAM* | A/G | 0.8 | 0.15 | -0.023 |
| LDL | rs11076175* | 16 | 57006378 | *CETP* | A/G | 0.16 | 0.23 | -0.021 |
| HDL | rs13326165 | 3 | 52532118 | *STAB1* | A/G | 0.89 | 0.19 | -0.022 |
| HDL | rs643531 | 9 | 15296034 | *TTC39B* | C/A | 0.95 | 0.09 | 0.043 |
| HDL | rs2167079 | 11 | 47270255 | *ACP2* | C/T | 0.48 | 0.09 | 0.019 |
| HDL | rs3184504 | 12 | 111884608 | *SH2B3* | T/C | 0.92 | 0.99 | 0 |
| TC | rs7515577 | 1 | 93009438 | *EVI5* | C/A | 0.96 | 0.7 | -0.011 |
| TC | rs2351524 | 2 | 203880992 | *NBEAL1* | T/C | 0.98 | 0.39 | -0.032 |
| TC | rs2255141 | 10 | 113933886 | *GPAM* | A/G | 0.8 | 0.05 | -0.028 |
| TG | rs4841132 | 8 | 9183596 | *NECAP2* | A/G | 0.9 | 0.44 | -0.016 |

Even when there are associations in the same region in two ancestry groups, it is possible that they are driven by different causal variants, as previously seen ^49^. To assess the extent of sharing of causal variants between ancestries at previously reported loci with evidence of transferability, we applied trans-ancestry colocalisation for G&H with UKBB EUR samples as the reference. We found evidence for the most extensive sharing of causal variants for transferable lipid loci: total cholesterol (61% of loci had significant colocalisation), followed by TG (56%), HDL-C (48%) and LDL-C (47%) (**Table 1**). For BMI we found evidence for sharing of causal variants for only 26% of transferable loci assessed (**Table 1** and **Table S9**). Causal variants in major lipid loci such as *PCSK9* were among variants that were consistently not shared (p_JLIM_>0.05) between the two populations (**Figure S6** and **Table S9**).

### Variable transferability of polygenic scores

Polygenic scores (PGSs) for CAD have been shown to have predictive value over risk scores based on clinical factors alone ^10,11,42,50–54^. To assess the transferability of PGSs for cardiometabolic traits derived from EUR populations into BPB individuals, we compared predictive performance in G&H to that in EUR individuals from eMERGE. We quantified predictive accuracy using the “incremental AUC” statistic for CAD and the “incremental R^2^” statistic for continuous risk factor traits; these are the gain in AUC or R^2^ when adding the PGS to the regression of phenotype on the baseline covariates (sex, age and genetic PCs).

We first evaluated the previously published PGSs from the PGS Catalog (**Table S5**). The PGSs for risk factors were developed using data from primarily EUR individuals, and the CAD PGSs that proved to have the best performance in G&H and eMERGE were two different scores optimised in SAS ^42^ and EUR samples ^53^, respectively. PGSs for all traits assessed were significant predictors in G&H (**Table S5, Figure 3A**). For prediction in G&H, the incremental R^2^ for BP was low (∼1.8%), but it was higher for lipids and BMI, ranging from 3.9% to 6.7%. Relative accuracy of PGS in eMERGE and G&H, determined by the ratio of incremental AUC or R^2^, was close to 1 for HDL-C, TG, SBP and DBP, and lower for CAD (42%, 95% CI: 30%–59%) and BMI (78%, 95% CI: 68%–88%; **Figure 3B**). Amongst the risk factors, prediction of LDL-C had the lowest relative accuracy (66%, 95% CI: 53%–79%), probably due to the fact that we did not adjust for statin usage since medication data were not available in eMERGE, and BPB individuals were more likely to be treated with statins ^55^. Incremental R^2^ for the PGS for LDL-C increased from 3.9% (3.3%–4.5%) to 6.2% (5.3%– 7.1%) when using statin-adjusted LDL-C in G&H (**Table S5, Figure 3A**), although the heritability was not significantly different (**Figure 2A**). In a sensitivity analysis, the relative accuracy of the CAD PRS in eMERGE *versus* G&H was consistent when defining CAD based on diagnostic codes only, rather than with the inclusion of procedure codes in the G&H definition (**Table S5**).

**Figure 3.**
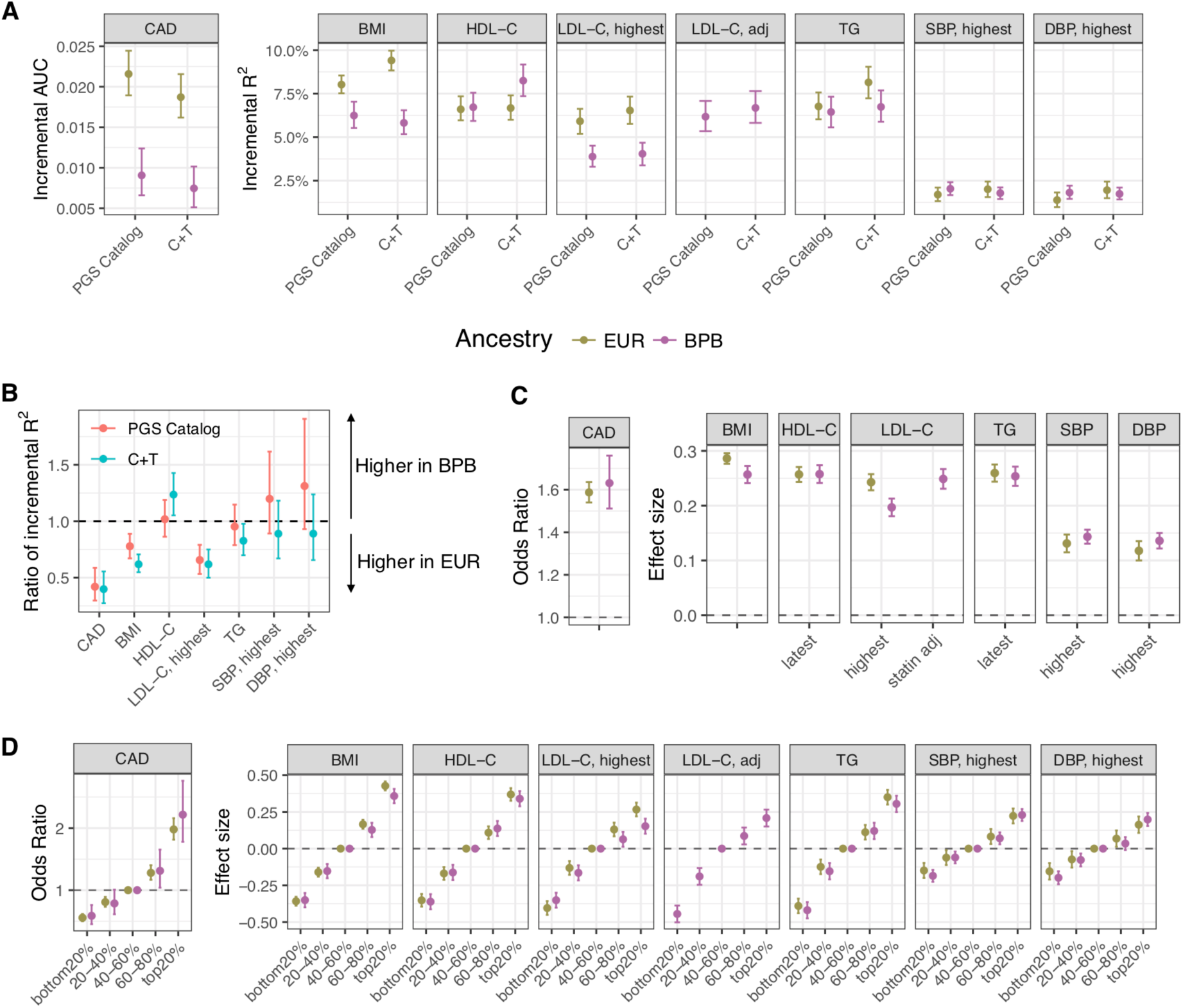
Comparison of the predictive accuracy of polygenic scores in people of British Pakistani and Bangladeshi versus European ancestry. **A**. Predictive accuracy of polygenic scores (PGSs) for cardiometabolic traits in British Pakistani and Bangladeshi (BPB) individuals from G&H (purple) and European-ancestry (EUR) individuals from eMERGE (green). Incremental AUC was calculated for coronary artery disease (CAD), and incremental R^2^ was calculated for its continuous risk factors. Error bars indicate 95% confidence intervals (CIs) estimated by bootstrap resampling of samples. The highest measurements for low-density lipoprotein cholesterol (LDL-C), systolic blood pressure (SBP), and diastolic blood pressure (DBP) are compared between eMERGE and G&H, and statin-adjusted LDL-C data are also shown for G&H. **B**. Relative accuracy of PGSs (i.e. the ratio of incremental AUC for CAD or incremental R^2^ for risk factors estimated in G&H to that in eMERGE) for PGS Catalog scores (red) and C+T scores (blue). Error bars represent 95% bootstrap CIs. Panel **C** and **D** show the effect sizes of PGSs from the PGS Catalog. **C**. The odds ratio per standard deviation (SD) of PGS is shown for CAD on the left panel, and the differences in phenotypic SD per SD of PGS are shown for quantitative traits on the right panel. **D**. The odds ratio for CAD comparing the four quintiles to the middle quintile (40–60%) is shown on the left panel. Quintiles are determined in controls. The differences in phenotypic SD compared to the reference quintile are shown on the right panel. Error bars show 95% CIs estimated using the standard error in **C** and **D**.

To assess whether the performance of PGS based on EUR GWAS could be improved in BPB, we next constructed PGS using the clumping and P-value thresholding (C+T) method and optimised them separately within G&H and eMERGE. The numbers of SNPs in the best C+T PGSs are similar between eMERGE and G&H, and PGSs for lipids contained fewer SNPs (194 to 454) than other traits (>20,000; **Table S10, Figure S7**). C+T PGSs and PGSs from the PGS Catalog showed similar performance in G&H across traits, although they were optimised in different ancestry populations (BPB and primarily EUR, respectively; **Figure 3A)**. For BMI, triglycerides and HDL-C, we observed slightly larger differences in predictive accuracies between G&H and eMERGE for C+T PGSs than observed with the PGS Catalog scores (**Figure 3B**).

We then assessed whether PGS methods that account for ancestry differences improved predictive accuracy in G&H. PGSs were constructed using a meta-score strategy ^41^, combining the EUR-derived PGS (described above) and that from UKBB SAS samples. The improvement in accuracy was modest (5–11%) (**Figure S8**). This may be due to the low sample sizes in the UKBB SAS GWASs.

### Modest improvement in CAD risk prediction by adding PGS to clinical risk score

A CAD PGS derived from EUR GWAS summary statistics and tuned in SAS individuals from UKBB^42^ (PGS000296 in the PGS Catalog), showed the highest predictive accuracy in BPB individuals in G&H. This score had an OR per SD of 1.63 (95% CI: 1.51–1.76) and incremental AUC of 0.009 (95% CI: 0.006–0.012; **Table S5**). Individuals in the top quintile of PGS were predicted to have a 2.2-fold increase (95% CI: 1.78–2.76) in disease risk relative to the middle quintile (quintiles were determined in controls; **Figure 3D**). We investigated the additional predictive power of PGS on top of established clinical risk factors for CAD, and the net reclassification improvement (NRI) achieved by adding the PGS to a clinical risk score.

To calculate the clinical risk score, we used the QRISK3 algorithm to estimate 10-year risk of cardiovascular disease at a baseline time point, selected so that the participants in G&H had about 10 years of follow-up. QRISK3 was a strong predictor of CAD events and had a concordance index (C-index) of 0.843 (95% CI: 0.828–0.858; **Figure 4A, Table S11**). Consistent with previous findings in EUR individuals^10^, the CAD PGS was uncorrelated with QRISK3 (Pearson’s correlation coefficient r=-0.0056 and p-value=0.62). We followed Riveros-Mckay *et al*.^10^ to construct an integrated score combining QRISK3 and the CAD PGS. The integrated score had a non-significant improvement in the C-index (0.853, 95% CI: 0.838– 0.867). However, compared with QRISK3 alone, the integrated score showed significant improvement in reclassification (categorical NRI: 3.9%; 95% CI: 0.9%–7.0%) using a 10-year risk threshold of 10% based on the threshold for preventive intervention with statin treatment recommended by National Institute for Health and Care Excellence ^56^. The integrated score reclassified 3.2% of the population as high risk and 2.5% as low risk (**Table S11**). This improvement was mostly driven by the enhanced identification of CAD cases in people at 25– 54 years old (NRI in cases being 7.0% *versus* NRI in controls being -1.2%), and of controls in people at 55–84 years old (NRI in cases being 0.0% versus NRI in controls being 6.8%) (**Figure 4B, Table S11**). The QRISK3 classified most (91.4%) of the individuals at 55–84 years old as high risk. Using the integrated score, 7.6% of the individuals older than 55 years were down-classified from high to low risk (**Table S11**). Using continuous NRI, the integrated score showed significant improvement (27.0%; 95% CI: 17.7%–36.2%) and similar trends in age groups (**Figure S9, Table S11**).

**Figure 4.**
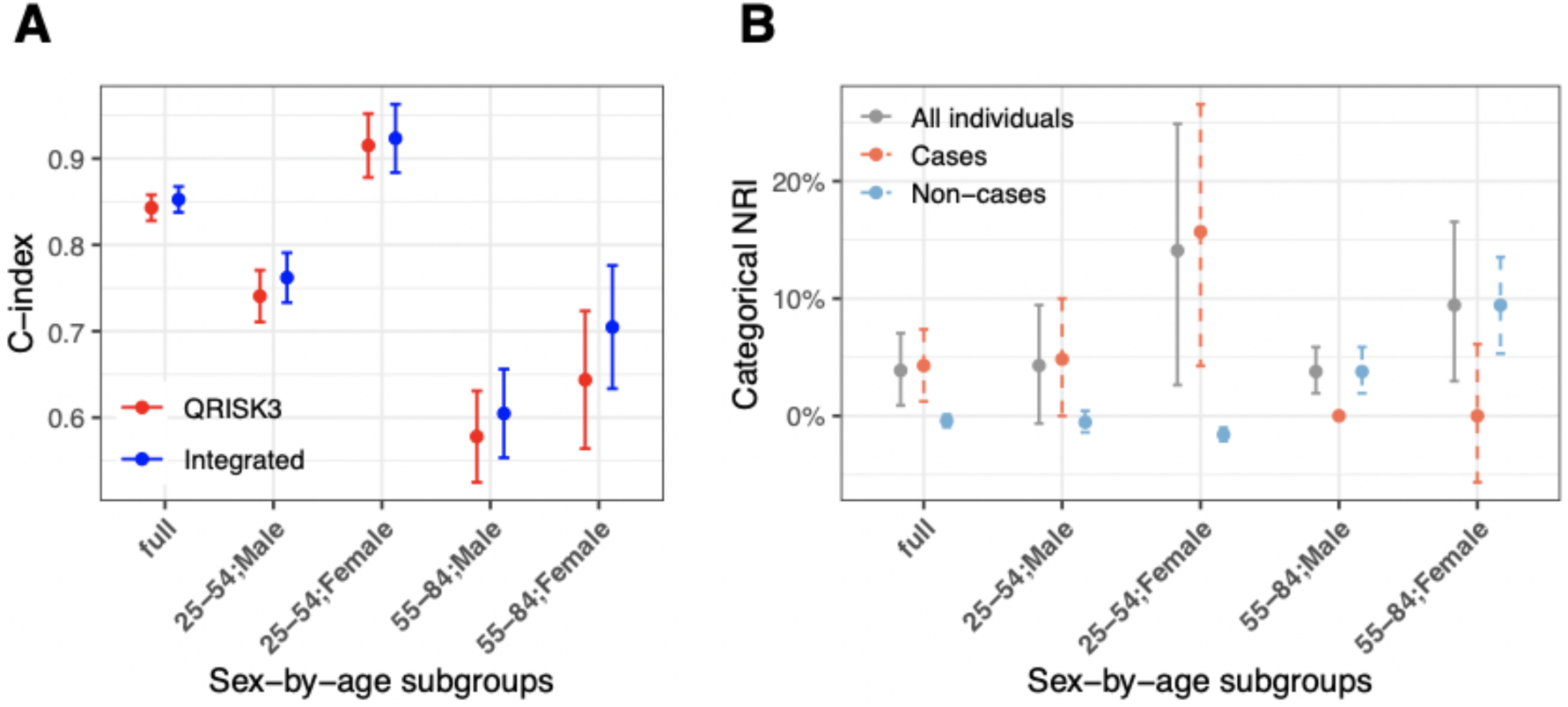
Model discrimination and net reclassification index for coronary artery disease with addition of a polygenic score to QRISK3. **A**. The concordance index (C-index) of QRISK3 (red) and an integrated score that combines QRISK3 and a polygenic score (PGS) for CAD (blue) in all British Pakistani and Bangladeshi individuals from G&H as well as in age-by-gender subgroups. The error bars represent 95% confidence intervals (CIs) estimated using the standard error. **B**. Categorical net reclassification index (NRI) for the integrated score compared to QRISK3 in all samples as well as in age-by-gender subgroups. NRI in cases (red) and controls (blue) are also shown. The error bars indicate 95% CIs estimated using the bootstrap method.

### Causal effects of CAD risk factors largely consistent across ancestries

We carried out two-sample Mendelian randomisation (MR) analyses to assess the causal effects of the risk factors on CAD in G&H and compared findings with EUR samples from eMERGE. For G&H, we used transferable loci as genetic instruments to benefit from the precision of large EUR discovery GWAS whilst ensuring only valid instruments are used. In eMERGE, causal effects for BMI, BP and lipids, except TG, were statistically significant (**Figure 5**). Consistent with this, we found that higher BMI (OR=1.73, p-value=0.01), higher LDL-C (OR=1.55 p-value=4×10^−4^) and lower HDL-C levels (OR=0.75, p-value=8×10^−3^) were causally associated with increased risk of CAD in G&H. The OR for LDL-C was larger than the one in eMERGE (OR=1.15) although with overlapping confidence intervals (CI: 1.03-1.29 in eMERGE, CI: 1.22-2.00 in G&H). The effects for SBP and DBP were not statistically significant in G&H. However, both had relatively small numbers of loci as instruments and confidence intervals of the effect estimates were wide.

**Figure 5.**
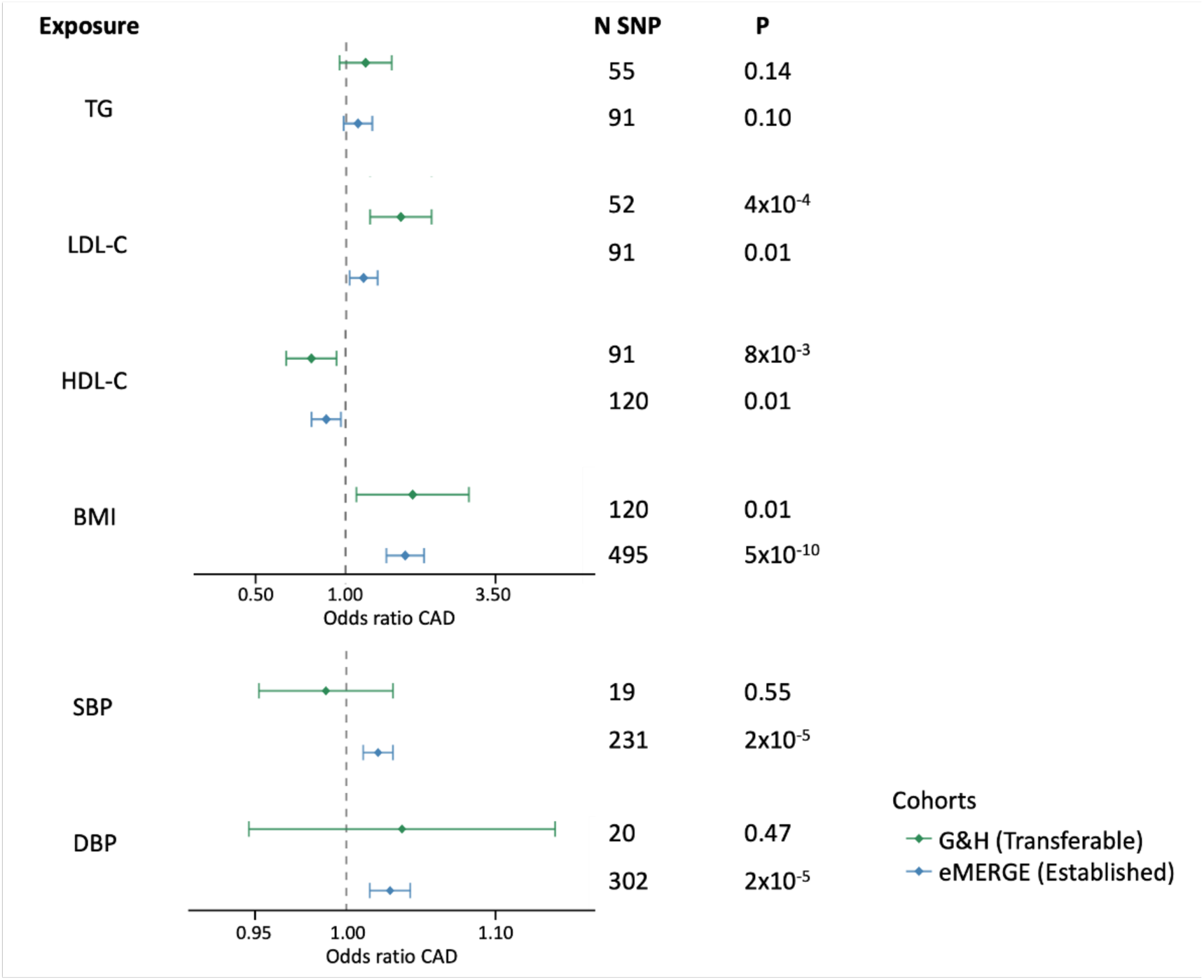
Estimates of causal effects of risk factors on coronary artery disease in European (eMERGE) and British South Asian (G&H) ancestry individuals. Two-sample Mendelian randomisation (MR) estimates for the causal effects are presented based on genetic instrument variables identified from EUR discovery GWAS for each risk factor. All independent genome-wide significant loci were used as instruments for eMERGE and only the transferable loci for G&H. Effect estimates are presented as odds ratios with 95% confidence intervals per standard deviation increase in the reported unit of the trait: triglycerides (TG), systolic blood pressure (SBP), low-density lipoprotein cholesterol (LDL-C), high-density lipoprotein cholesterol (HDL-C), diastolic blood pressure (DBP), body mass index (BMI). The p-value (P) and number of single nucleotide polymorphism instruments (N SNPs) included in the MR analysis are shown for each exposure.

We also assessed different strategies for instrument selection in G&H, such as using all loci associated at genome-wide significance in EUR GWAS for the risk factors (**Figure S11**). When following the standard approach of using an independent ancestry-matched sample (UKBB SAS) to derive the instruments, an insufficient number of genome-wide significant instruments (p<5×10^−8^) were identified (**Figure S10**). To address this, we also tested a less stringent p-value threshold (p<5×10^−5^) for selecting instruments. For the lipid biomarkers, the results were consistent regardless of which loci were chosen as instruments (**Figure S11**). However, the association of BMI with CAD was significant only for transferable loci (**Figure S11**).

We found evidence of heterogeneity between causal estimates based on Cochran’s Q statistic for DBP when using the established loci as instruments (p-value=0.04), LDL-C when using the UKBB SAS-ascertained loci (p-value=0.02) and HDL-C for transferable loci (p-value=1×10^−3^). However, the results of the weighted median and weighted mode models were consistent with those obtained by the inverse-variance weighted MR model (**Table S12**).

## Discussion

We conducted the first study to systematically assess the transferability of genetic loci and PGSs for cardiometabolic traits in SAS individuals with real-world clinical data, using ∼22,000 individuals from the G&H cohort. For lipids and blood pressure, we found evidence that causal genetic variants at known loci and beyond are widely shared with EUR. The prediction accuracy of PGSs derived from EUR GWASs for these traits was similar between G&H and EUR samples. However, the predictive performance of BMI and CAD PGS was reduced by 22 and 58%, respectively (for the PGS Catalog scores), in G&H, and CAD also had fewer transferable loci. A CAD PGS optimised for South Asians nonetheless yielded an appreciable improvement in risk reclassification when combined with the QRISK3 clinical risk score.

Other genetic studies of CAD and related traits that have evaluated reproducibility of established loci in SAS populations have either been limited by small sample sizes or have restricted their comparisons to the index SNP identified in the GWAS, which does not take LD into account ^34–36^. A recent study compared genetic determinants of >200 lipid metabolites in 5,000 South Asians from Pakistan and 13,000 Europeans and found high overlap in the detected associations ^57^. Using a new method, our paper goes further by empirically demonstrating that, in most cases where loci do not replicate, it is due to lack of power. These findings suggest that, in large part, the genes and pathways that influence risk of CAD are shared between these ancestrally divergent populations. One surprising finding was that the major LDL-C locus at *CETP* was not associated with this biomarker in G&H but exhibited pleiotropic effects particularly on HDL-C. Abnormalities in *CETP* are linked to accelerated atherosclerosis and might play an important role in increasing risk in SAS ^58^.

Of those previously reported cardiometabolic loci that contained variants significantly associated in G&H, 30–74% did not show evidence of shared causal variants. This suggests that, although the genes and pathways are likely to be shared between ancestral groups, there is heterogeneity with respect to the causal alleles. BMI had the lowest proportion of transferable loci with shared causal variants as well as lower transferability of the PGS in G&H and a genetic correlation significantly lower than one. SAS individuals are known to have higher visceral fat at the same BMI compared to EUR individuals in Western countries ^59,60^. Consistent with this, the causal effect of BMI was significant only when using the transferable loci as instruments in the Mendelian randomisation analysis. Visceral adiposity is a strong risk factor for cardiometabolic diseases, independent of total fat mass; these findings warrant further study and may suggest that BMI may not be an optimal biomarker of adiposity in SAS^61^.

Mendelian randomisation has emerged as a powerful tool to explore the causal effects of risk factors on disease outcomes. Statistical power can be the limiting factor when extending these analyses to non-EUR populations because independent ancestry-matched GWAS for risk factors of interest may not be sufficiently large. To increase power to estimate the causal effects of risk factor traits on CAD in BPB, we used genetic instruments derived from large EUR GWAS. Some of the loci may be invalid instruments for other populations. However, restricting the established loci to the ones that were transferable in this population successfully addressed this issue for BMI and shows promise as a new approach for trans-ancestry Mendelian randomisation. An assumption that requires further study is whether the effect sizes of transferable loci are the same for each ancestry group.

We observed variable levels of PGS transferability from EUR into BPB individuals for the cardiometabolic traits that were investigated in this work, with relative accuracy in G&H versus eMERGE ranging from 131% for DBP to 42% for CAD. Consistent with previous studies ^37,62^, PGSs for HDL-C and triglycerides had similar predictive accuracy between the two ancestry groups. We explored the factors that may impact relative accuracy of PGSs. Based on a recently proposed theory, relative accuracy is proportional to the product of the trans-ethnic genetic correlation and the ratio of heritability estimates ^7^. We considered the effect on the relative accuracy of the trans-ethnic genetic correlation, ratio of heritability estimates in G&H versus eMERGE, as well as the product of the previous two terms. However, none of them showed a significant association with the relative PGS performance (**Figure S12**). This may be because the theory was derived for PGSs based on genome-wide significant SNPs (whereas our PGSs include many SNPs with less significant p-values), and because the relative accuracy also depends on differences in allele frequencies and LD patterns at these SNPs between populations, which we have not factored in and may differ between traits.

Based on findings in lipid traits, the Global Lipids Genetics Consortium recently claimed that GWASs with high enough sample sizes could lead to PGSs with equally high accuracy across ancestry populations, even if the GWASs were conducted in predominantly EUR samples ^62^. However, we do not fully agree that this claim can be generalised beyond lipid traits, since it depends on the extent to which the causal variants are shared across ancestry groups. For example, the accuracy of C+T PGS for BMI decreased by 38% in G&H, whereas that for TG decreased by only 17% and that for HDL-C did not decrease, although the sample size of the input GWAS for BMI was much larger than that for lipids (about 700,000 versus 300,000; **Table S4**). This is likely due to the relatively lower fraction of shared causal variants (26%) at transferable loci for BMI and the relatively lower genetic correlation (significantly lower than 1 for BMI while close to 1 for lipids), which will not be ameliorated with larger sample sizes of Europeans.

Several groups have shown improvements in PGS performance in non-Europeans when incorporating summary statistics from ancestry-matched samples ^41,63^. Incorporating UKBB SAS GWAS data in meta-PGSs proposed by Marquez-Luna et al. ^41,63^ did not show large improvement in G&H. A likely reason is the limited sample size of the SAS samples in UKBB for some of the traits. Larger samples of SAS individuals are needed to examine if ancestry-matched GWAS data can improve prediction accuracy over and above what would be expected from the increased sample size. For traits for which the causal variants are shared, there is more to be gained from more powerful EUR GWASs, even without adding samples of the target ancestry. However, increasing diversity in GWASs will greatly improve the resolution of fine-mapping and the power to identify the causal variants by leveraging the LD differences across ancestries ^64^.

We assessed the clinical value of the PGS for CAD on top of the traditional clinical risk factors captured in the QRISK3 algorithm. Similar work has been done previously in research cohorts ^9–12^; our study represents an important addition since it captures the noise with which QRISK3 is actually measured within a real-world clinical setting (as opposed to using comprehensive measures taken for research purposes), which may affect performance of integrated risk models combining these factors with PGSs. We note that only about 4% of the ∼8 million individuals used for developing QRISK3 were of South Asian ancestry ^26^, and the weights for each conventional risk factor might not be optimal for SAS individuals. QRISK3 was developed to predict cardiovascular disease (CVD), which is a composite outcome of CAD and stroke. However, our analysis focused on CAD, which is an important component of CVD and the main focus in GWASs and genetic prediction studies. The PGS for CAD developed by Wang *et al*. showed robust association with CAD in G&H, with a similar OR per SD in PGS (1.63, 95% CI: 1.51–1.76) as in their study (1.60, 95% CI: 1.32–1.94) ^42^. The integrated score combining PGS and QRISK3 showed significant reclassification improvement against QRISK3 alone (NRI 3.9% (95% CI: 0.9–7.0%)). Previous studies in UKBB EUR samples reported similar improvement, with NRI estimates of 3.5% (95% CI: 2.4–4.5%) ^10^ and 3.7% (95% CI: 3.0–4.4%) ^9^ in two different analyses using CAD as the outcome. However, these NRI estimates are probably inflated by using UKBB samples that are healthier than the general UK population without recalibrating risk to a primary care setting^11^. In G&H, the PGS improved identification of high-risk individuals in people younger than 55 years, and correctly down-classified low-risk individuals in people older than 55 years, both of which are important in a clinical setting. We anticipate that, like EUR individuals ^9–11^, the British Pakistani and Bangladeshi community (and potentially other SAS populations) would also benefit from the use of integrating PGS in primary prevention settings.

Our study has several limitations. Firstly, due to the limited sample size in each age-by-sex subgroup, we could not recalibrate risk prediction models in G&H to what would be expected in an unbiased primary care setting ^11^. Secondly, while the G&H cohort has enabled us to assess the potential utility of genetics in an under-represented population using data from electronic records, each of the cohorts examined here is unique. Differences in ascertainment (including the age distribution) and clinical measurements within different cohorts and healthcare systems may have impacted the genetic associations. Ideally future studies would compare populations with different ancestries collected in the same real-world healthcare setting, but with sufficient sample sizes in each ancestry group to enable well-powered comparisons. The BioMe biobank in New York contains individuals from multiple ancestries with linked EHR data, but the number of self-reported SAS individuals is very limited (N=622)^65^.

In conclusion, our work provides the first comprehensive assessment of the transferability of cardiometabolic loci to a non-EUR population and its impact on two key applications of genetics, causal inference and risk prediction. Our protocol and our new approach for transferability can serve as methodological standards in this developing field. We have shown high transferability of GWAS loci across several cardiometabolic traits between EUR and BPB populations. The transferability of PGSs is trait-specific. Our results suggested there would be clinical value in adding PGS to conventional risk factors in the prediction of CAD in primary care settings to improve the more efficient use of preventive interventions, such as lipid-lowering medications. Our investigation contributes to the increasing representation of individuals of non-European ancestry and lower socio-economic status in research studies, which we hope will help to decrease health disparities.

## Data Availability

Genes & Heath imputed genotype data will be available on EGA (study accession number: EGAS00001005373). Researchers wishing to access phenotype data should apply to the G&H Executive (https://www.genesandhealth.org/research/scientists-using-genes-health-scientific-research). GWAS summary statistics generated in Genes & Health are available at https://www.genesandhealth.org/research/scientific-data-downloads. Publicly available GWAS summary statistics that were used in this study are provided in Supplementary Table S4.

## Acknowledgements

We thank Social Action for Health, Centre of The Cell, members of our Community Advisory Group, and staff who have recruited and collected data from volunteers. We thank the NIHR National Biosample Centre (UK Biocentre), the Social Genetic & Developmental Psychiatry Centre (King’s College London), Wellcome Sanger Institute, and Broad Institute for sample processing, genotyping, sequencing and variant annotation. We thank Barts Health NHS Trust, NHS Clinical Commissioning Groups (Hackney, Waltham Forest, Tower Hamlets, Newham), East London NHS Foundation Trust, Bradford Teaching Hospitals NHS Foundation Trust, and Public Health England (especially David Wyllie) for GDPR-compliant data sharing. We also thank Sally Hull and Martin Sharp from the primary care data team at QMUL for their help in estimating population prevalence of CAD. Most of all we thank all of the volunteers participating in Genes & Health.

## Supplementary Materials

Supplementary Figures 1–12

Supplementary Tables 1–12

