## Supplementary Figures for "Transferability of genetic loci and polygenic scores for cardiometabolic traits in British Pakistanis and Bangladeshis"

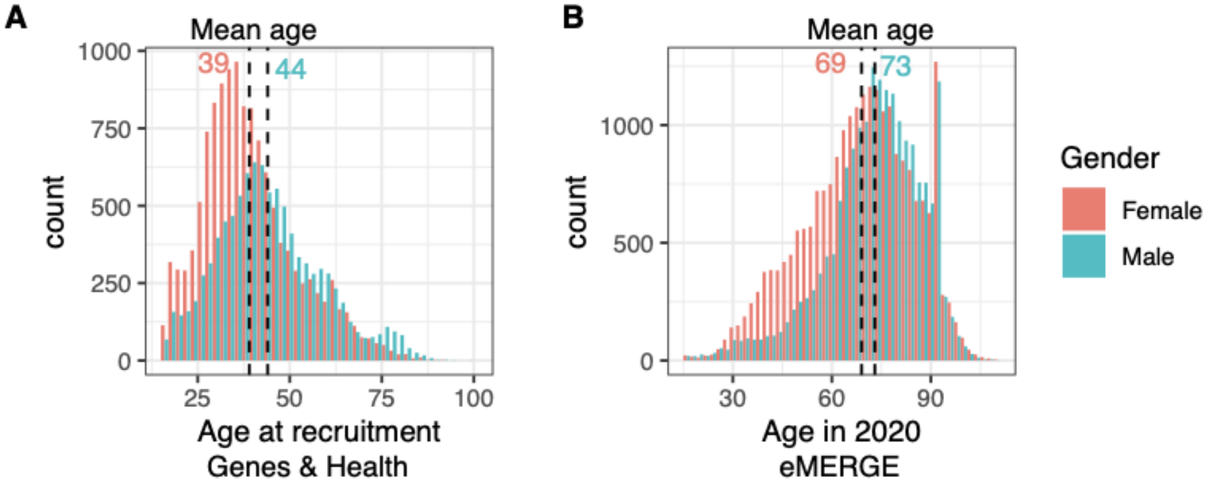


**Supplementary Figure 1. Age distributions of G&H (A) and eMERGE (B)**. Red indicates female participants and blue indicates male participants. Vertical dashed lines indicate the average age. In G&H, 56.5% of the 22,490 individuals with electronic health record data are female, with the mean age 39.4 (standard deviation, SD: 13.1) years old for women and 44.3 (SD: 14.3) for men. In eMERGE, 54.5% of the 42,802 individuals are female, with the mean age 69.1 (SD: 16.7) and 73.1 (SD: 14.7) for women and men, respectively.

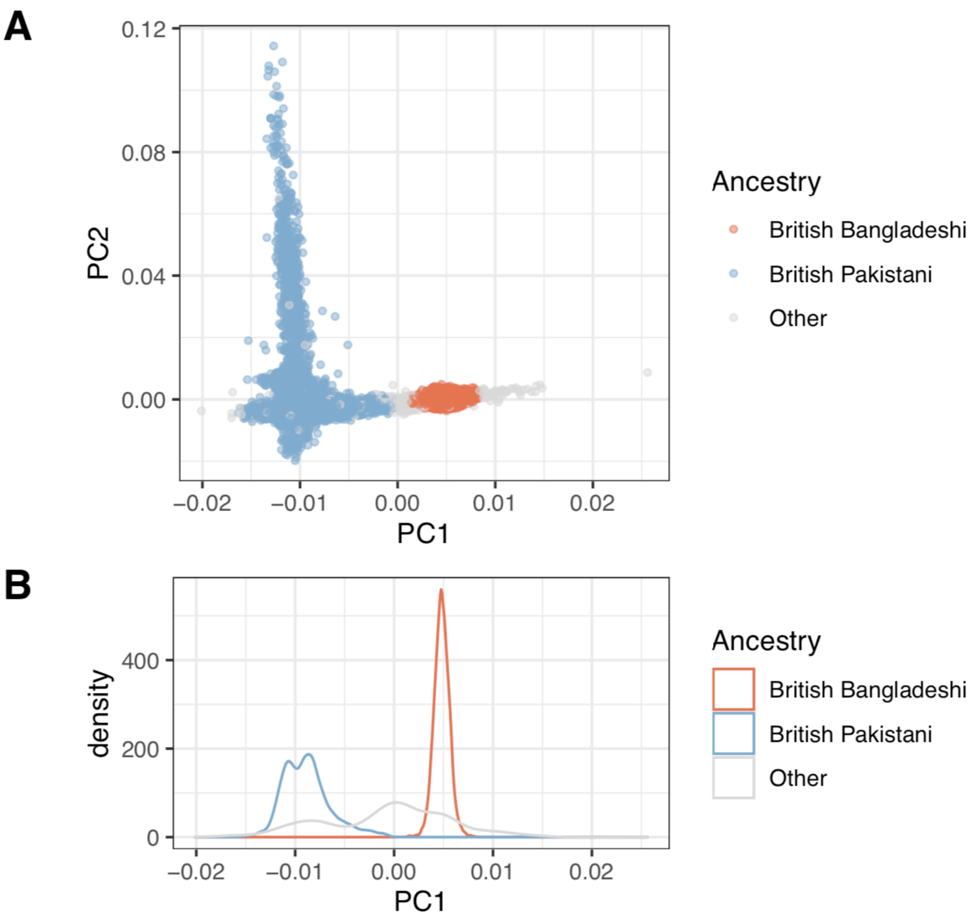


**Supplementary Figure 2. Principal component analysis (PCA) of 28,022 genotyped individuals from the Genes & Health (G&H) cohort. A**. PC1 and PC2 for all samples. Red indicates self-declared British Bangladeshi samples (N=17,721) and blue represents self-declared British Pakistani samples (N=9,694). We excluded samples who self-reported as coming from other ethnic groups ("Other") or who did not report ethnicity information, as well as genetically-inferred outliers (those with PC1 further than +/- 3 standard deviations from the mean of PC1 for the individuals who self-reported as coming from that group). These samples are in grey (N=607). **B**. Density plot for PC1, which clearly differentiates self-declared British Bangladeshis from self-declared British Pakistanis.

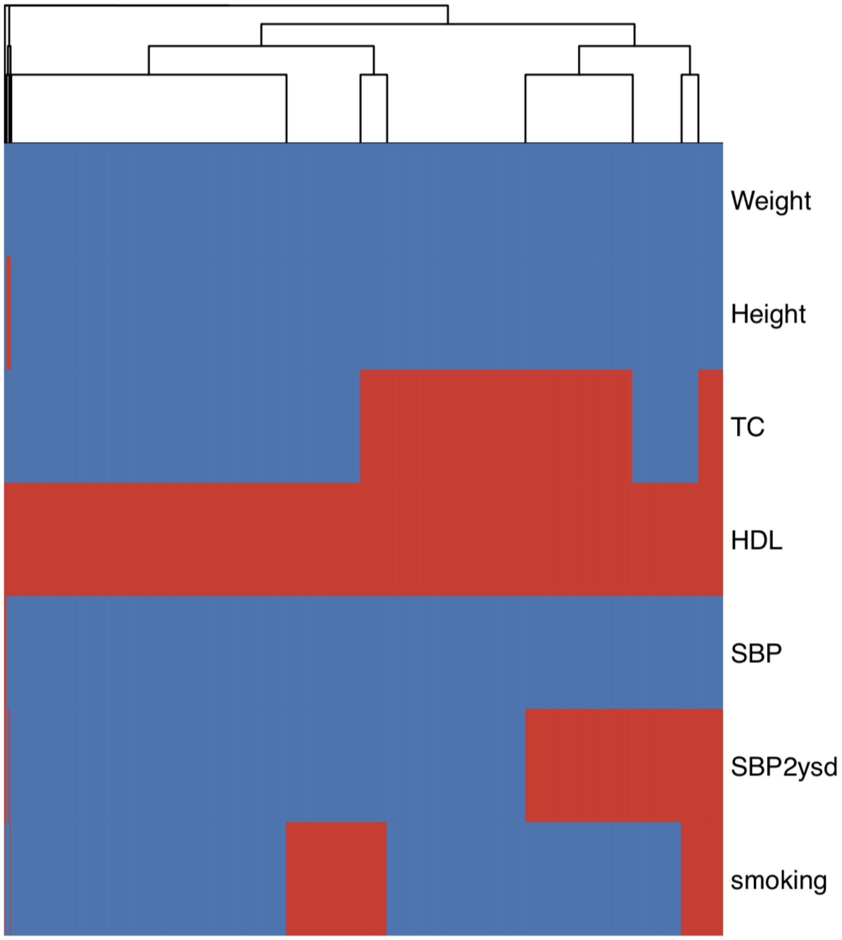


**Supplementary Figure 3. Pattern of missingness for continuous variables in the QRISK3 algorithm**. Columns represent individuals (N=9,477, before removing relatives) and rows represent variables (TC: total cholesterol; HDL: high-density lipoprotein cholesterol; SBP: systolic blood pressure; SBP2ysd: standard deviation of SBP measurements within 2 years). Missing data are in red.

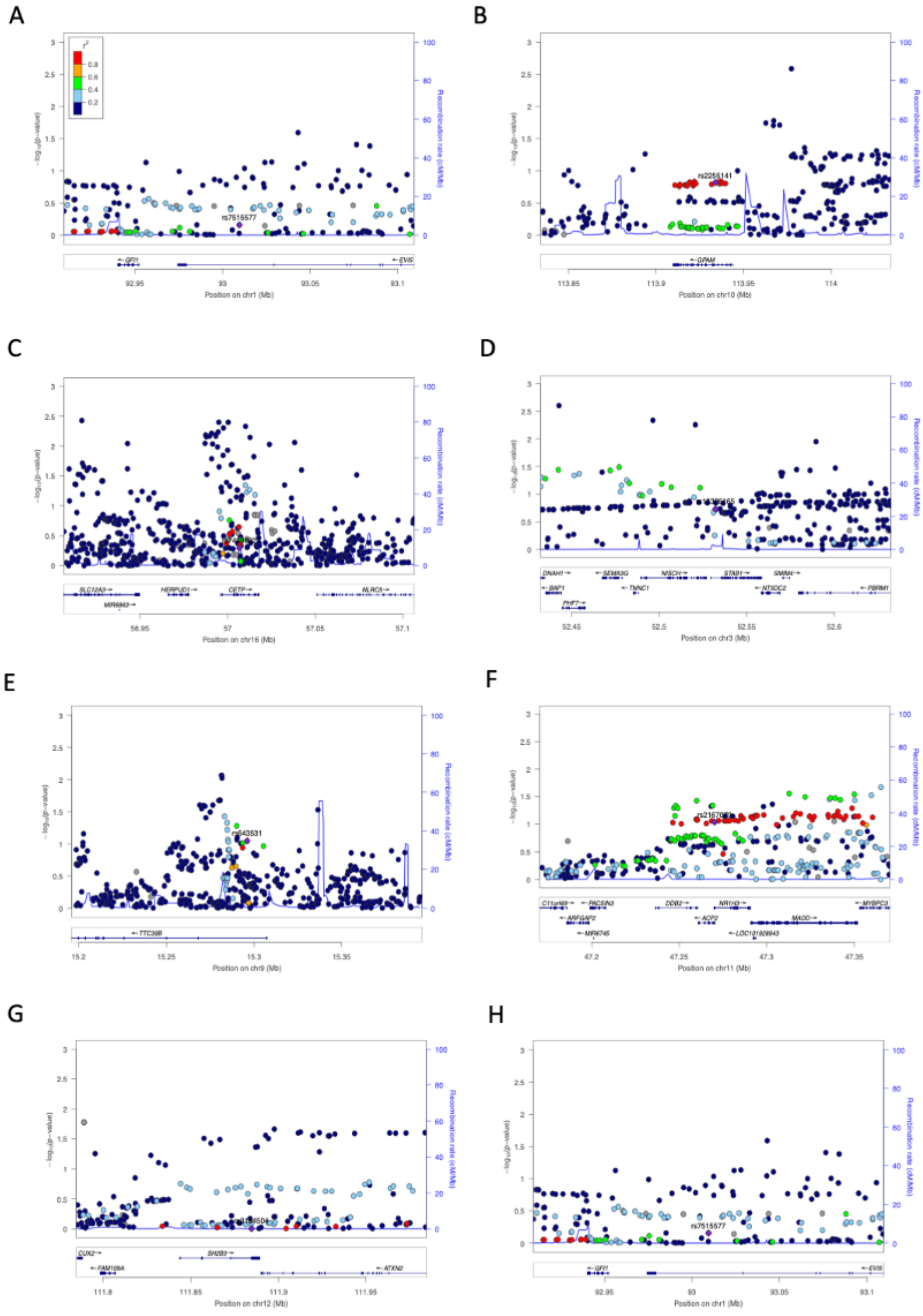
A
B
C
D
E
F
G
H
rs7515577
rs2255141
GFI1
EVI5
GPAM
SLC12A3
HERPUD1
CETP
NLRC5
MIR6863
DNAH1
SEMA3G
NISCH
STAB1
SMIM4
BAP1
TNNC1
NT5DC2
PBRM1
PHF7
TTC39B
rs643531
C11orf49
PACSIN3
DDB2
NR1H3
MYBPC3
ARFGAP2
ACP2
MADD
MIR6745
LOC101928943
CUX2
SH2B3
FAM109A
ATXN2
Position on chr1 (Mb)
Position on chr10 (Mb)
Position on chr16 (Mb)
Position on chr3 (Mb)
Position on chr9 (Mb)
Position on chr11 (Mb)
Position on chr12 (Mb)
Recombination rate (cM/Mb)

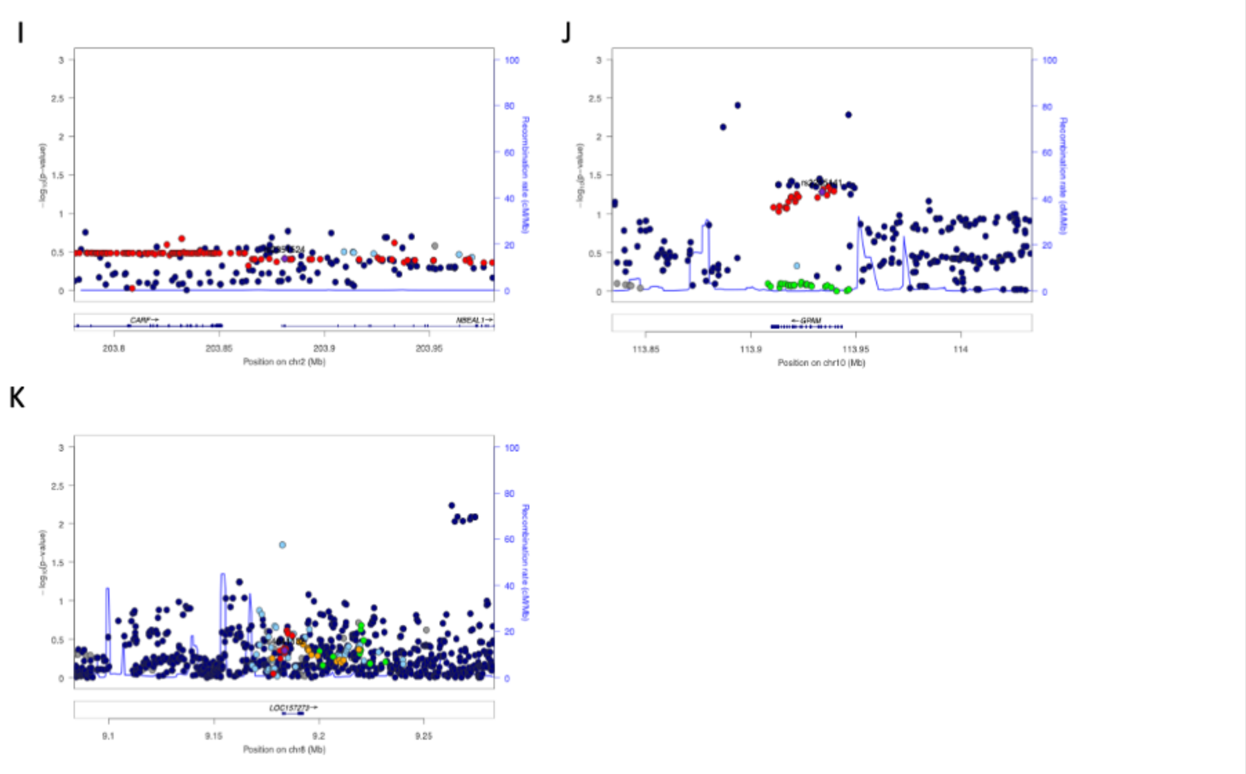


**Supplementary Figure. 4. Regional association plots for non-transferable loci in G&H (Credible set p > 0.05 and no variant within 50kb of locus with p < 1x10$^{-3}$).** **A.** LDL-C, *EVI5* locus (rs7515577). **B.** LDL-C, *GPAM* locus (rs2255141). **C.** LDL-C, *CETP* locus (rs7499892). **D**. HDL-C STAB1 locus (rs13326165). **E.** HDL-C, *TTC39B* locus (rs643531). **F.** HDL-C, *ACP2* locus (rs2167079). **G.**HDL-C, *SH2B3* locus (rs3184504). **H.** Total Cholesterol, *EVI5* locus (rs7515577). **I.** Total Cholesterol, *NBEAL1* locus (rs2351524). **J.** Total Cholesterol, *GPAM* locus (rs2255141). **K.** Triglycerides, *NECAP2* locus (rs4841132). Colour of the points corresponds to the strength of linkage disequilibrium ($r^2$) of each potential causal variant (in brackets) identified in EUR ancestry labelled and coloured in purple.

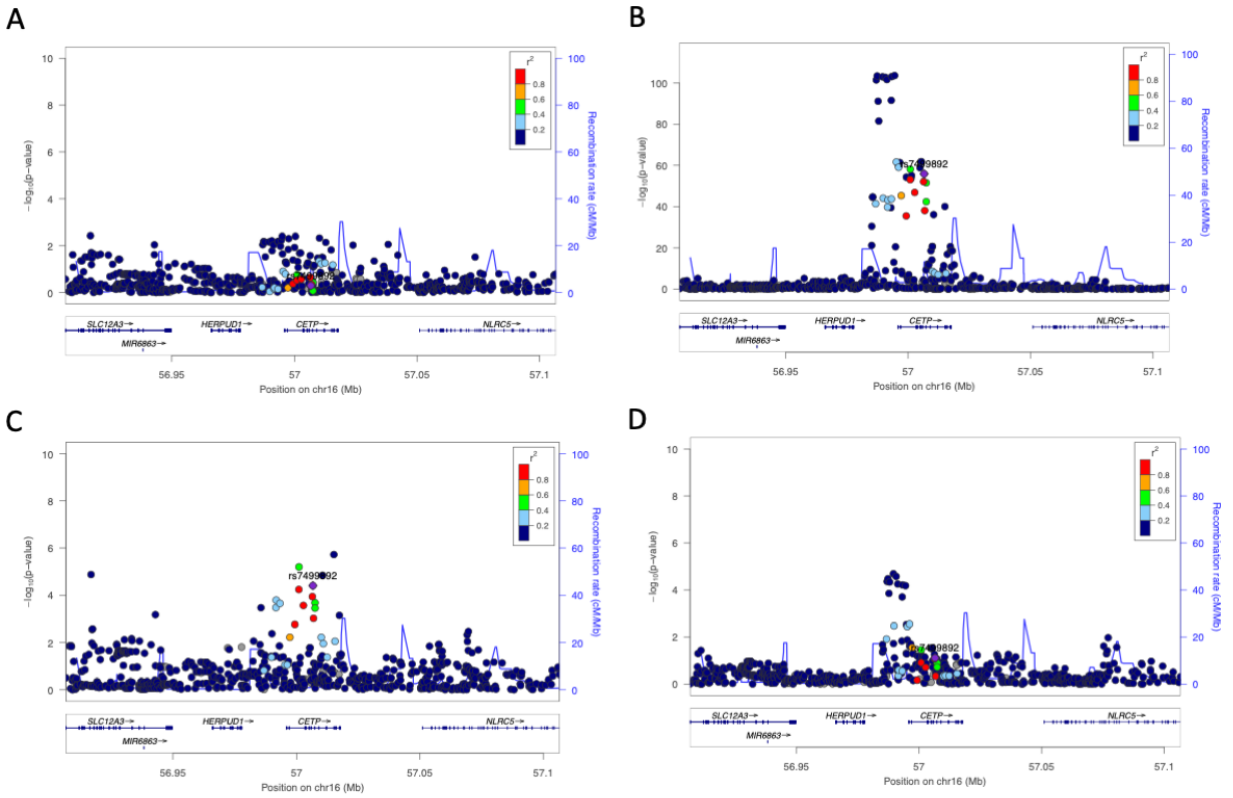


**Supplementary Figure. 5. Regional association plots for *CETP* locus across lipid traits in G&H**. **A.** LDL-C. **B.** HDL-C. **C.** Total Cholesterol. **D.** Triglycerides. Colour of the points corresponds to the strength of linkage disequilibrium ($r^2$) of potential causal variant (rs7499892) identified in EUR ancestry, labelled and coloured in purple.

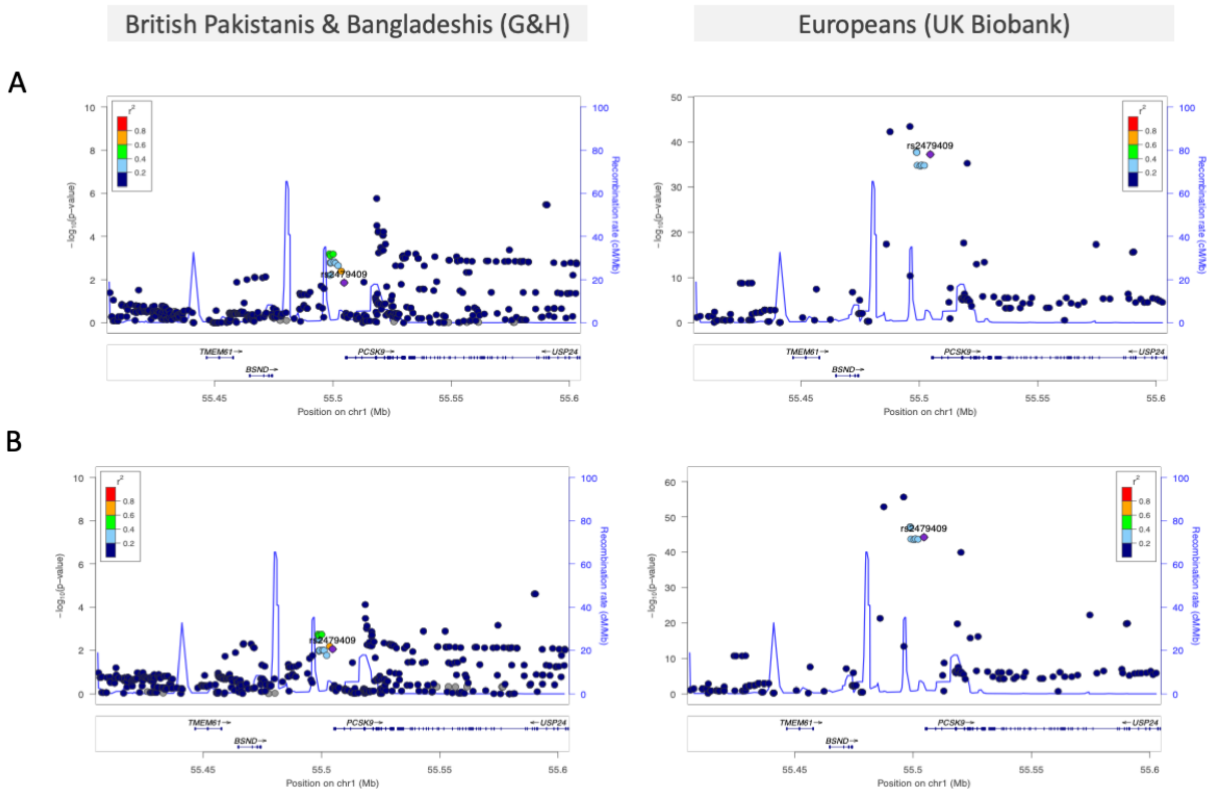


**Supplementary Figure. 6. Regional association plots for major lipid locus, *PCSK9* with unshared causal variant rs2479409 ($p_{JLIM}$>0.05) between British Pakistanis and Bangladeshis (G&H) and Europeans (UK Biobank). A.** LDL-C. **B.** Total Cholesterol. Colour of the points corresponds to the strength of linkage disequilibrium ($r^2$) of each potential causal variant (in brackets) identified in EUR ancestry, labelled and coloured in purple.

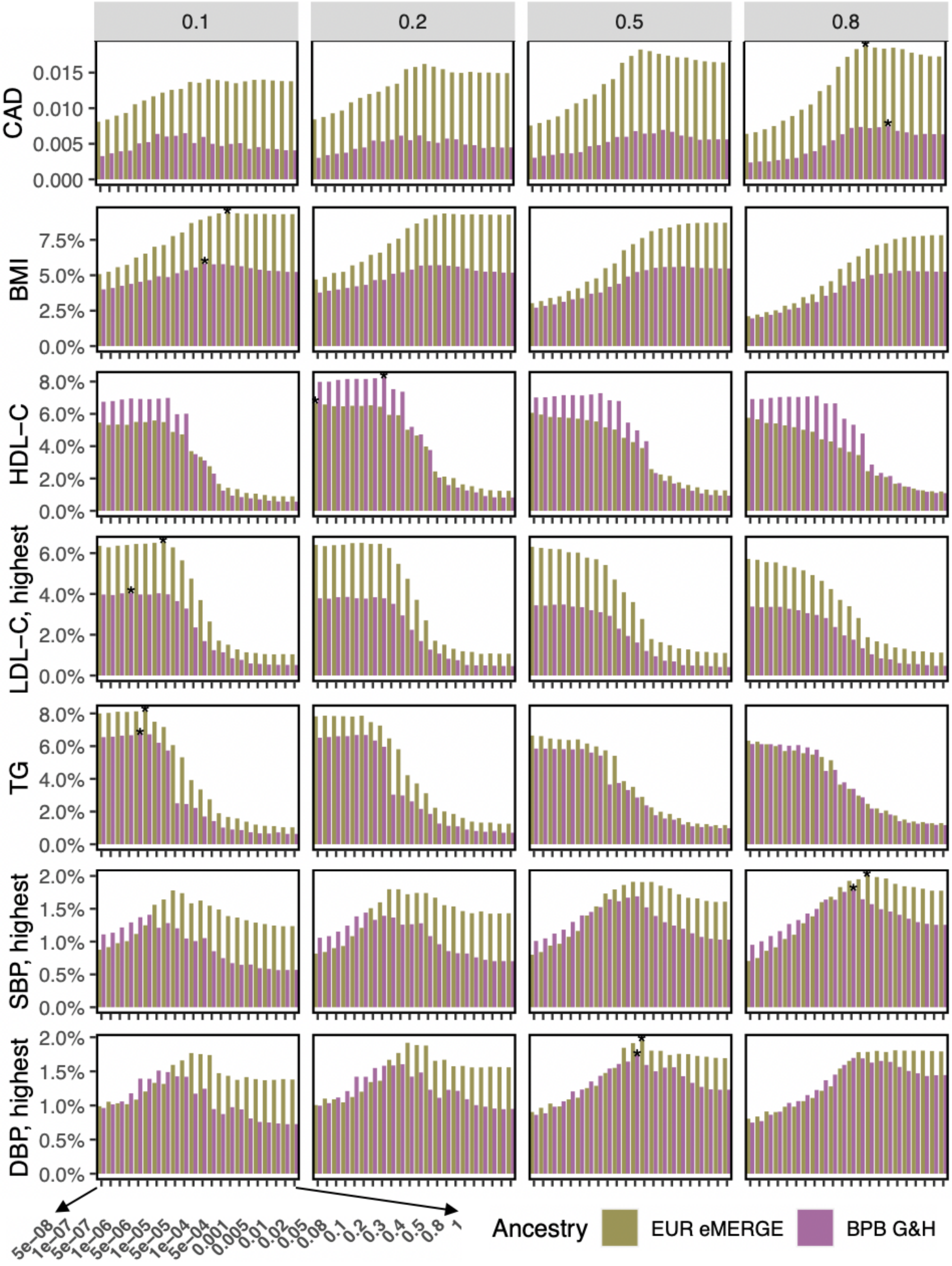


**Supplementary Figure 7. Predictive performance of polygenic scores (PGSs) across LD $r^2$ and p-value thresholds.** Incremental AUC is shown for CAD and incremental $R^2$ is shown for the continuous risk factors. Purple indicates the performance in G&H and green in eMERGE. Bars represent PGSs constructed using combinations of various p-values (on x-axes) and LD clumping $r^2$ thresholds (in the four columns). PGSs are sorted so that those on the right contain more SNPs. Asterisks indicate the reported PGSs with the highest accuracy per trait.

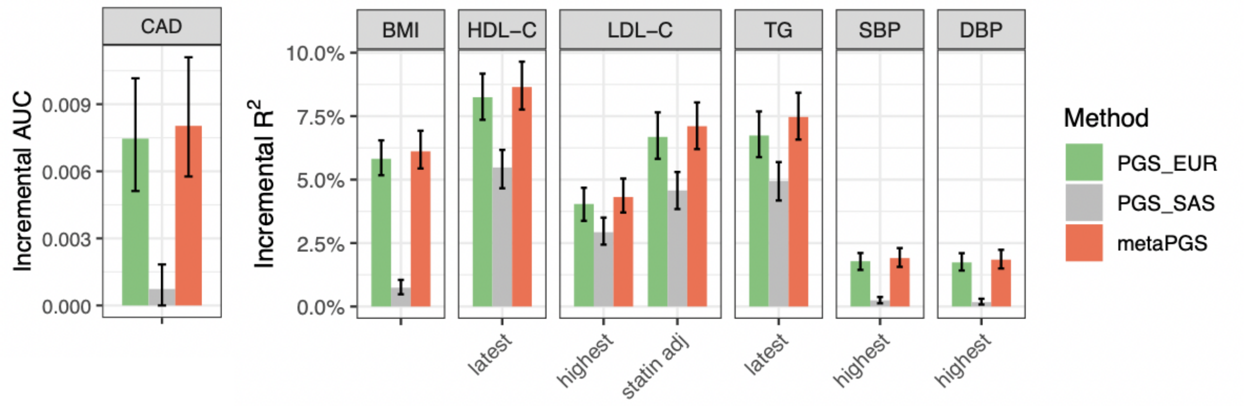


**Supplementary Figure 8. Predictive performance of polygenic scores (PGSs) that incorporate GWAS data from UK Biobank (UKBB) South Asian-ancestry individuals.** Incremental AUC was calculated for CAD, and incremental $R^2$ was calculated for its continuous risk factors. Colours indicate clumping and p-value thresholding PGSs that are constructed using different GWAS training data (green: GWAS statistics from European studies; grey: GWAS statistics from South Asian-ancestry samples from UKBB; red: meta-PGS, a linear combination of the previous two PGSs). Error bars indicate 95% confidence intervals estimated by bootstrap resampling of samples. Improvement in accuracy comparing the meta-PGSs (red) with the C+T PGS derived from European studies (green) is 8% for CAD, 5% for BMI, 5% for HDL-C, 7% for the highest LDL-C, 6% for the statin-adjusted LDL-C, 11% for TG, 7% for SBP, and 6% for DBP.

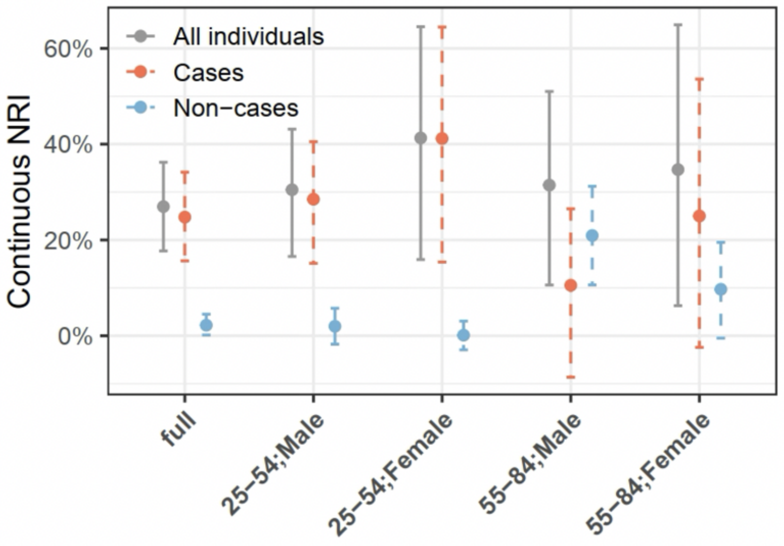


**Supplementary Figure 9. Continuous net reclassification index (NRI) for the integrated score compared to QRISK3 in all samples and age-by-gender subgroups.** Continuous NRI in cases (red) and non-cases (blue) are shown. The error bars indicate 95% CIs estimated using the bootstrap method.

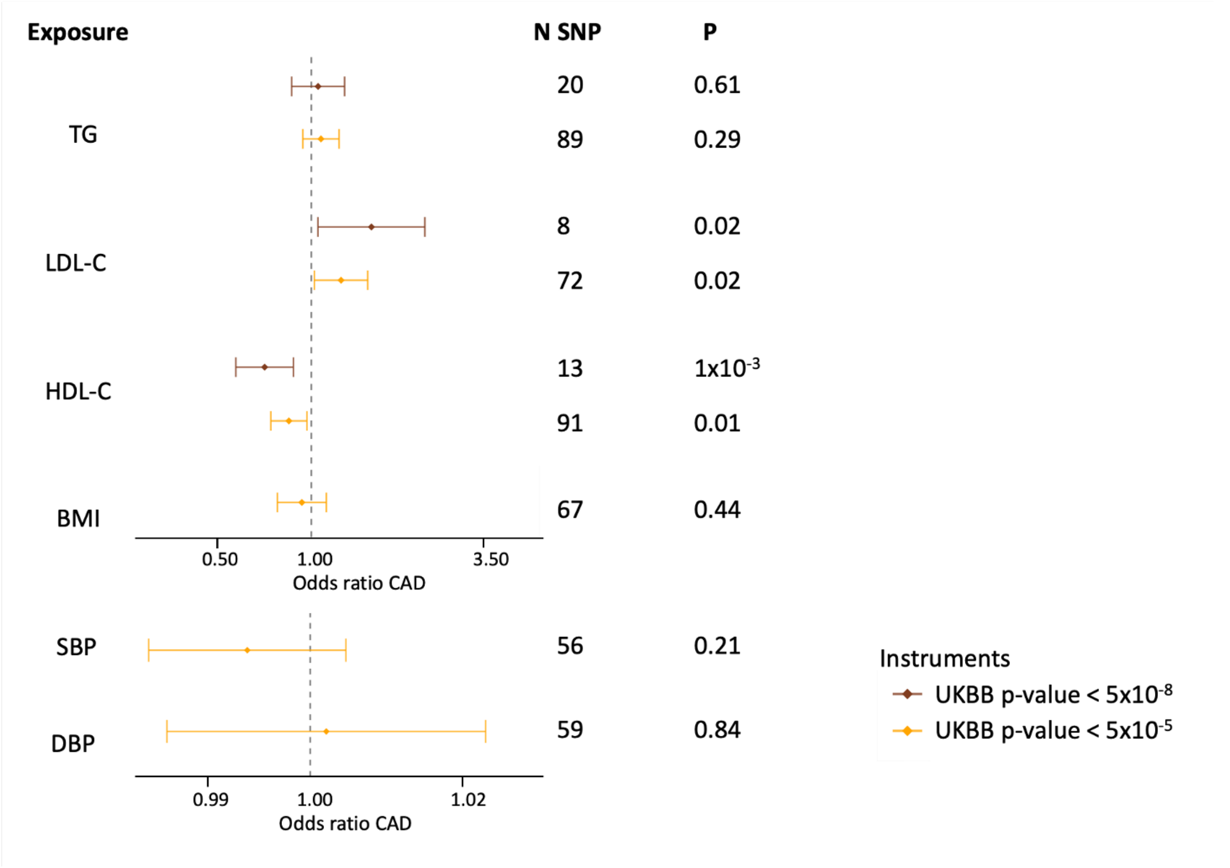


**Supplementary Figure 10. Mendelian randomisation (MR) estimates of the causal effects of risk factors on coronary artery diseases (CAD) in G&H using loci from ancestry-matched discovery GWAS as instruments.** Association of risk factors with CAD was assessed for instruments selected from UKBB SAS at two p-value thresholds: $p<5x10^{-5}$ and $p<5x10^{-8}$. Effect estimates are presented as odds ratios with 95% confidence intervals per standard deviation increase in the reported unit of the trait: triglycerides (TG), systolic blood pressure (SBP), low-density lipoprotein cholesterol (LDL-C), high-density lipoprotein cholesterol (HDL-C), diastolic blood pressure (DBP), body mass index (BMI). The p-value (P) and number of SNP instruments (N SNP) included in the MR analysis are shown for each exposure.

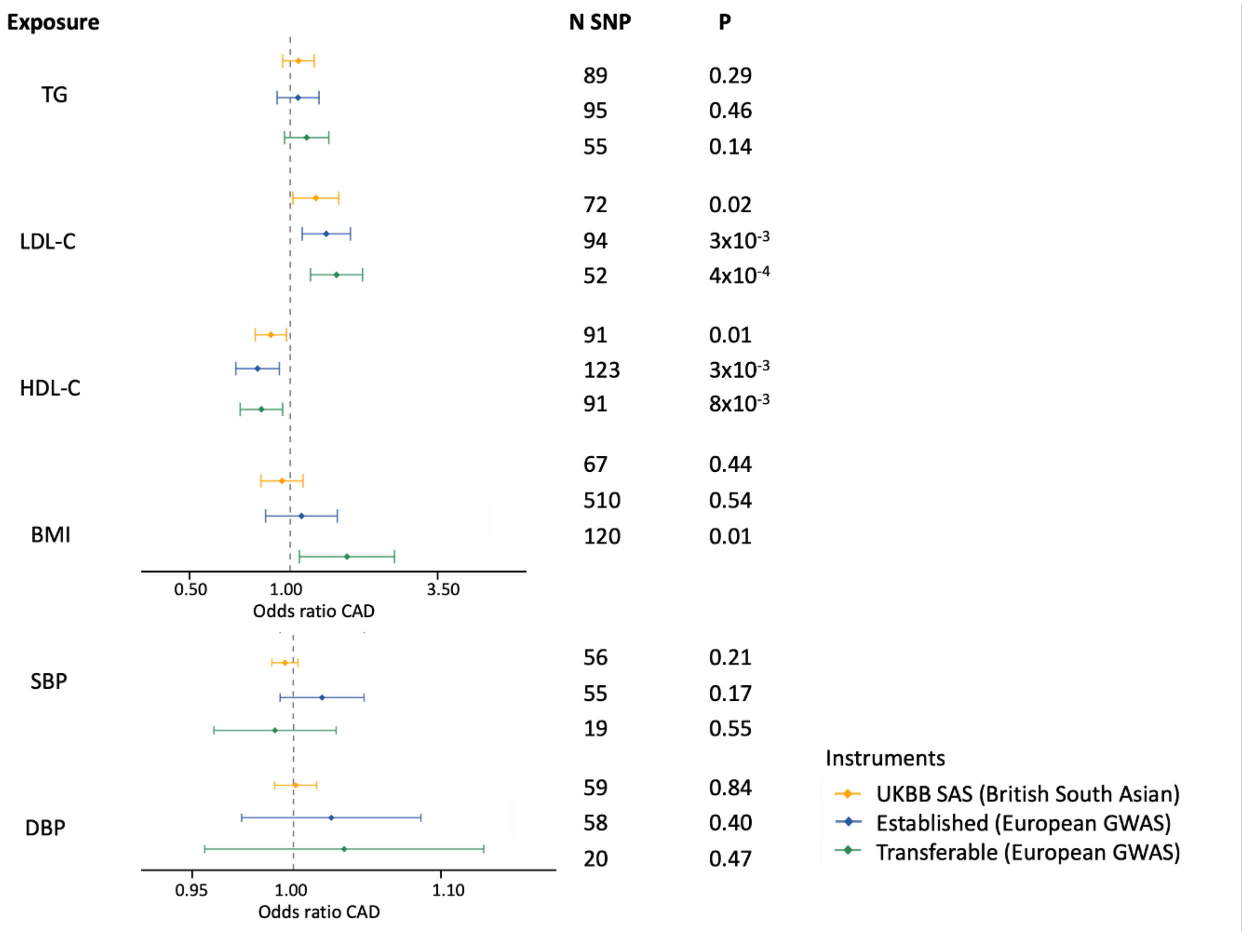


**Supplementary Figure 11. Mendelian randomisation estimates for risk factors on coronary artery disease using different strategies for instrument selection.** Two-sample Mendelian Randomisation (MR) using coronary artery disease risk (CAD) in G&H as the outcome. Genetic instrumental variables for the risk factors were selected based on different strategies: loci associated at $p<1x10^{-5}$ in an ancestry-matched GWAS (UKBB SAS), all genome-wide significant loci from the largest EUR GWAS, and the subset of these loci that were transferable to SAS. Effect estimates are presented as odds ratios with 95% confidence intervals per standard deviation increase in the reported unit of the trait: triglycerides (TG), systolic blood pressure (SBP), low-density lipoprotein cholesterol (LDL-C), high-density lipoprotein cholesterol (HDL-C), diastolic blood pressure (DBP), body mass index (BMI). The p-value (P) and number of single nucleotide polymorphism instruments (N SNP) included in the MR analysis are shown for each exposure.

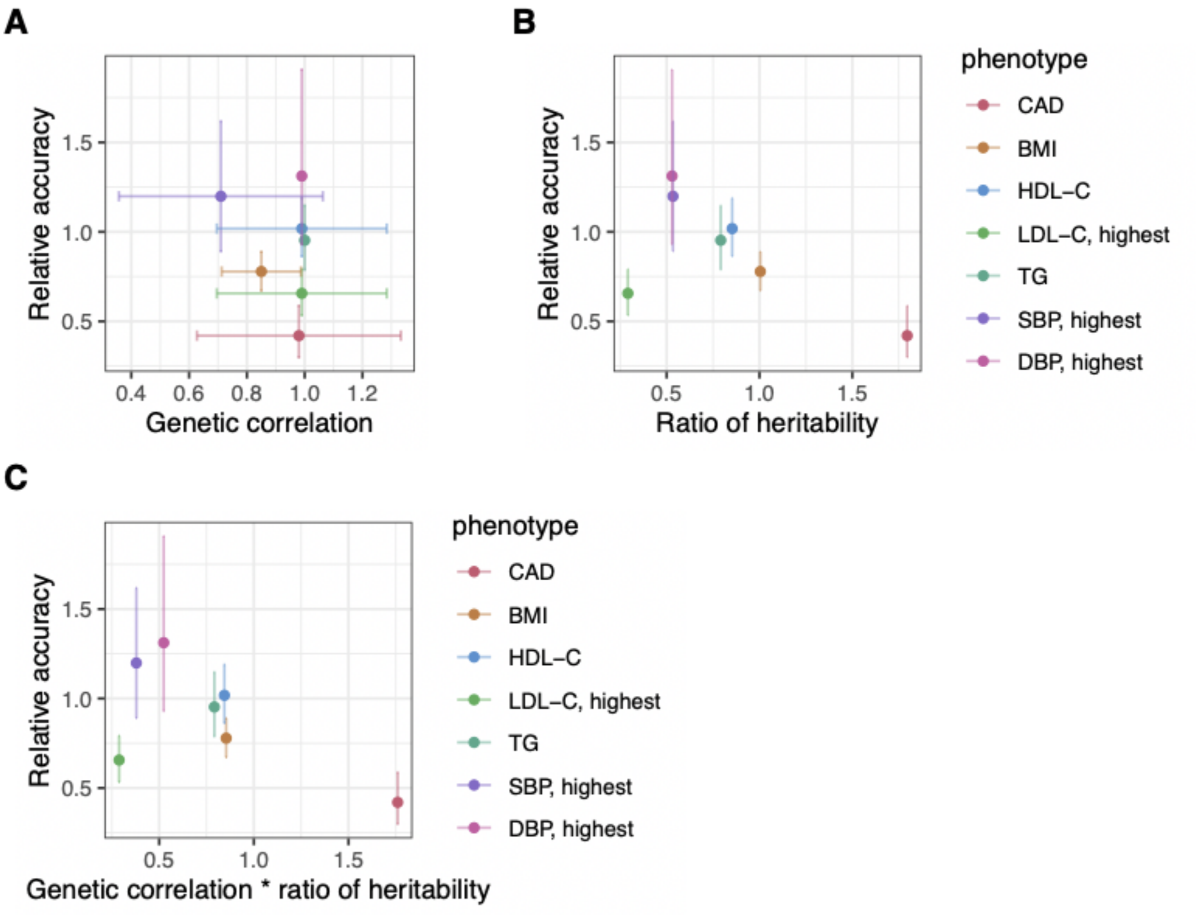


**Supplementary Figure 12. Relationship between the relative accuracy of PGSs and trans-ethnic genetic correlation and heritability estimates.** We used PGSs from the PGS Catalog. We show the relationship between relative accuracies of PGSs (i.e. the ratio of incremental AUC for CAD or incremental $R^2$ for risk factors estimated in G&H to that in eMERGE) on the y-axes versus the trans-ethnic genetic correlation (**A**), ratio of the heritability estimates in G&H over UKB (**B**), and the product of the previous two terms (**C**). None of the correlations is significant.
